# Automated bioinformatic pipeline for unbiased detection of tuberculosis transmission clusters: Real-time impact and retrospective insights

**DOI:** 10.64898/2026.03.16.26348245

**Authors:** Charlotte Genestet, Quentin Testard, Genna Ben-Hassen, Claire Bardel, Maxime Vallée, Chloé Bourg, Olivier Bahuaud, Brune Joannard, Caroline Tatai, Sophie Barabotti, Florence Ader, Cédric Dananché, Elisabeth Hodille, Oana Dumitrescu

## Abstract

**Background:** In low-burden countries such as France, whole-genome sequencing (WGS) is increasingly integrated into routine tuberculosis (TB) surveillance to improve case management and transmission monitoring. However, applying WGS to all TB cases generates large volumes of data, requiring automated tools for timely interpretation and outbreak response.

**Methods:** Since November 2016, all clinical *M. tuberculosis* isolates diagnosed in eight hospitals from three cities of Auvergne-Rhône-Alpes in France have undergone WGS. In July 2023, an automated pipeline for anti-TB drug resistance prediction and unbiased detection of transmission clusters based on SNP distances was implemented. Epidemiological, microbiological and clinical data were collected, with contact duration classified as household, frequent, or occasional. Index cases were stratified by their level of extra-household transmission (EHT), and statistical analyses were performed to identify associated factors.

**Findings:** Among 1,152 TB patients diagnosed between 2016 and 2025, 75 clusters involving 247 patients (21·4%) were identified. WGS reliably detected resistance to first-line anti-TB drugs, leveraging the WHO mutation catalogue. Routine WGS enabled real-time alerts for TB control centres, leading to expanded field investigations, including community spillover, nosocomial transmissions, and school outbreak. Classical indicators of contagiousness (smear results, cavitary disease) were not associated with EHT level. Instead, lower TB severity indices and longer duration of symptoms were linked to higher EHT level.

**Interpretation:** Systematic WGS supports timely identification of drug resistance and transmission events and provides new insights into contagiousness factors. The automated pipeline enables direct interpretation by clinical microbiologists, facilitating real-time public health action. In this study, we demonstrate how, with the appropriate pipeline, WGS offered a time- and cost-effective solution for routine TB management.

**Funding:** This work was supported by SHAPE-Med@Lyon, a French government grant managed by the French National Research Agency under the France 2030 program (reference ANR-22-EXES-0012).

**RESEARCH IN CONTEXT:** *Evidence before this study:* A systematic literature search was conducted using PubMed up to October 2025, with the following keywords: “tuberculosis” AND “whole genome sequencing” AND “contact tracing” OR “population based” OR “genomic surveillance” OR “investigation”. The review encompassed studies addressing the use of whole-genome sequencing (WGS) for tuberculosis (TB) surveillance, transmission cluster detection, and integration into routine epidemiological practice. Most published studies have focused on using WGS to retrospectively confirm or refute transmission events or to distinguish relapse from reinfection and therefore have not used genomic data to trigger real-time alerts to TB control centres. Even in the rare studies adopting a more proactive approach, the operational integration of WGS into routine diagnostic workflows is not described. No prospective study has provided a clear framework for the real-time use of WGS by clinical microbiologists to enable routine implementation and direct public health action.

*Added value of this study:* To our knowledge, this is the first study presenting the implementation of an automated WGS pipeline that enables unbiased detection of TB transmission clusters, based on single nucleotide polymorphism (SNP) distance, in routine surveillance across multiple hospitals. It shows how WGS data can be directly interpreted by clinical microbiologists for anti-TB drugs susceptibility prediction as well as facilitating real-time notification of TB control centres and immediate field investigations. By integrating genomic, epidemiological and clinical data, this comprehensive approach provides new insights into the factors associated with extra-household transmission and moves beyond the retrospective use of WGS, establishing its role as a proactive tool in TB diagnosis and public health practice.

*Implications of all the available evidence:* These findings highlight the need for national and international guidelines to evolve and support the widespread integration of WGS into routine TB management. The adoption of automated pipelines, such as the one presented herein, would not only enhance epidemiological surveillance but also streamline resistance detection and species identification, providing a time- and cost-effective all-in-one tool. Broad implementation of WGS-based approaches could significantly improve public health responses and the overall management of TB.

## INTRODUCTION

Tuberculosis (TB), caused by the *Mycobacterium tuberculosis* (Mtb) complex, remains a persistent public health challenge in Europe. For instance, countries of the European Union and European Economic Area (EU/EEA) reported a notification rate of approximately 8·6 per 100,000 population and France of 7·1 per 100,000 population in 2023.^1^ Furthermore, multidrug-resistant (MDR) TB accounted for on mean 4·3% reported cases among EU/EEA countries and 1·4% of TB cases in France in 2023.^1,2^ Given the clinical and public health implications of drug resistance (treatment complexity, costs, and outcomes), molecular and epidemiological surveillance through whole genome sequencing (WGS) in Europe has focused primarily on MDR-TB.^3,4^ This focus is essential for effective patient management and containment of resistant strains. Nevertheless, since MDR-TB represents a minority of TB cases in low-incidence settings, a broader surveillance is needed to enable earlier detection and more effective control of overall transmission. Applying WGS to all TB cases, however, generates a much larger volume of data, necessitating automated tools that enable direct interpretation by clinical microbiologists. While previous studies demonstrated the value of Mtb WGS to decipher transmission clusters or distinguish a reinfection from a relapse,^5–9^ none of them proposed an interface allowing direct analysis by clinical microbiologists and therefore real-time feedback to field investigators. Such tools are essential to facilitate real-time reporting to clinicians and to public health authorities responsible for implementing TB patient treatment, contact tracing and outbreak management. Herein, we present how an automated bioinformatic pipeline for identification of drug-resistance mutations and the unbiased detection of TB transmission clusters was integrated into routine TB surveillance. Based on real-time reporting of WGS results, this pipeline enabled timely initiation of the appropriate anti-TB treatment and as well as rapid identification of transmission events and more targeted interventions. Furthermore, by combining genomic, epidemiological and clinical data, this comprehensive approach has uncovered risk factors for extra-household transmission that differ from the classical indicators of TB contagiousness typically used in field investigations.

## METHODS

### Study population

Between January 2008 and October 2025, the mycobacteria laboratory at the university hospital of Lyon (Auvergne-Rhône-Alpes region of France), labelled French reference medical biology laboratory for next-generation sequencing (NGS) for TB, conducted a multicentre molecular epidemiology survey. Spoligotyping was performed on 7,563 Mtb isolates collected from all university hospitals in the Auvergne-Rhône-Alpes region and those diagnosed in the region by the private laboratory Biomnis, with approximately 400 to 450 isolates genotyped each year over a 18-year period.^10,11^ WGS was carried out since November 2016 on Mtb isolates diagnosed in eight hospitals across three cities (Lyon, Valence, and Bourg-en-Bresse), representing between 100 and 130 TB cases each year.^12^

The present multicentre study included all microbiologically-confirmed TB cases, with available Mtb WGS data, diagnosed between November 2016 and October 2025. To ensure optimal surveillance of MDR-TB, 25 MDR Mtb isolates identified between January 2010 and October 2016 were also included (appendix p 10). Phenotypic drug susceptibility testing was performed using the MGIT-960 SIRE kit (MGIT-960; Becton Dickinson Diagnostic Systems, Franklin Lakes, NJ, USA).^12^

### WGS and bioinformatic pipeline

Genomic DNA extraction, library preparation, short-read WGS using Illumina technology (San Diego, CA, USA) and bioinformatic analyses were performed as previously described (https://github.com/samlipworth/snpit).^11,13^ The reference genome coverage breadth was >92% with a mean depth of coverage >25x. The 2021 WHO catalogue of mutations in Mtb complex and their association with drug resistance was used as a database to detect antibiotic resistance.^13–15^

For unbiased detection of TB clusters based on SNP distance, once the 25x sequencing depth threshold was reached, VCF files generated by the pipeline’s variant calling module were filtered. Only SNPs supported by ≥5 reads and an allele frequency ≥0·80 were retained, while indels were excluded.^16^ The resulting filtered VCF was then converted into a FASTA consensus sequence using bcftools consensus (v1.15.1). The consensus sequence of each sample was integrated into a pre-existing database and a comprehensive multi-FASTA compilation file. The tool snp-dists (v0.8.2) was then employed to generate a SNP distance matrix for all strains from this compiled file (appendix p 11). For strain pairs separated by a distance ≤20 SNPs, a joint VCF was generated. Subsequently, Integrative Genomics Viewer (IGV)-reports (v1.7.0) was used, integrating the joint VCF and the respective BAM files, to facilitate manual curation. This critical step enabled the inspection of variants that may have fallen below quality thresholds or remained undetected by the initial variant calling module (appendix pp 12-13). The within-host Mtb micro-diversity passed from the index to the secondary case was excluded from pairwise SNP distance analyses and was instead leveraged as an indicator to refine reconstruction of transmission links.^17^

### Integration of WGS data into contact tracing investigations

WGS data were integrated into prospective TB surveillance from July 2023 through the automated generation of the distance matrix. Each week, clinical biologists performed WGS data analysis and reviewed the resulting distance matrix. When the pairwise SNP distance between two isolates was ≤20, the IGV-report was used for manual curation. For isolates with a SNP distance ≤12 after manual curation, potential laboratory contamination was assessed by verifying that samples had been processed in different batches at inoculation and sequenced in separate WGS runs. If laboratory contamination was excluded, a SNP distance ≤5 was interpreted as evidence of recent transmission, whereas a distance of 6-12 SNPs suggested older transmission or the presence of an intermediary case.^18^ In both cases a notification was sent to the TB control centre. When putative index and secondary cases had been hospitalised during the same prior period, the hospital infection control department was informed (figure 1).

**Figure 1:**
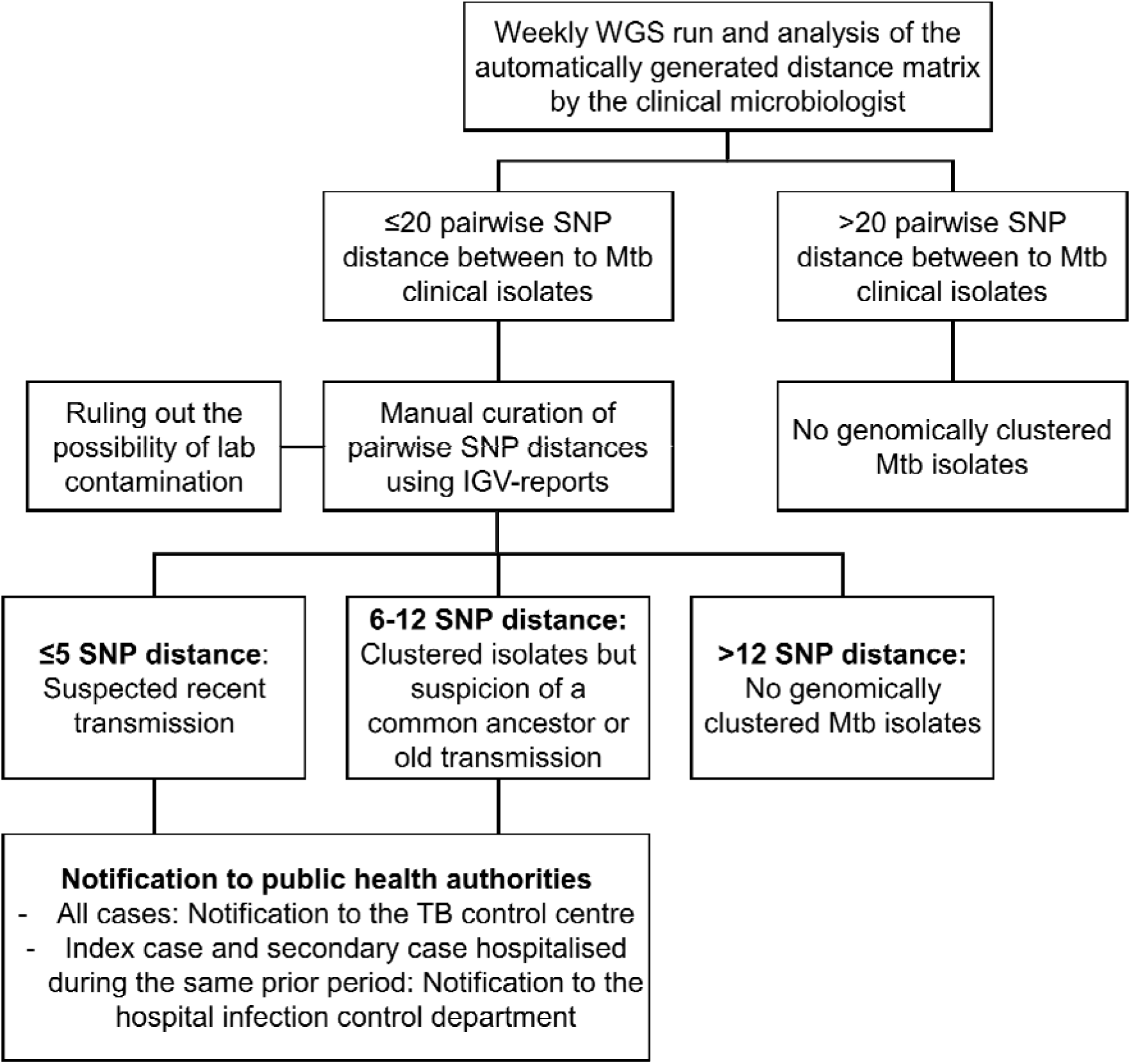
Decision tree based on WGS data. IGV: Integrative Genomics Viewer; Mtb: *Mycobacterium tuberculosis*; SNP: Single Nucleotide Polymorphims; TB: Tuberculosis; WGS: Whole-Genome Sequencing.

### Patient data collection

For all participants, microbiological data were recorded, including lineage and drug resistance profiles. For cases identified within transmission clusters, epidemiological data were documented, meaning contact duration between index and secondary cases classified as household, frequent (family, friends, colleagues, …), or occasional (<8 cumulative hours or not identified during field investigations). For index cases, detailed demographic and clinical information were recorded, meaning TB clinical presentation (including the Bandim TB score which considers five symptoms and five clinical findings, one point each),^19^ comorbidities, substance use, haematological markers and nutritional status (including the Malnutrition Universal Screening Tool [MUST] that considers unintentional weight loss, BMI, and anorexia).^19^ For secondary cases, data collection focused on pre_TB immune status. Individuals were considered immunocompromised if they met any of the following criteria: HIV coinfection, immunosuppressive treatment, or age >75 years. Based on the epidemiological data and pre-TB immune status of secondary cases, index cases were stratified into three ordered categories according to their level of extra-household transmission (EHT): exclusively within household (HH-restricted), to one immunocompetent secondary case outside of household (Low-EHT) and to two or more immunocompetent secondary cases outside of household (High-EHT).

### Ethics

All data were collected in a database in accordance with the decision 20-216 of the ethics committee of the Lyon University Hospital and French legislation in place at the time of the study (reference methodology MR-004 that covers the processing of personal data for purposes of study, evaluation, or research that does not involve the individual). Relevant approval regarding access to patient-identifiable information are granted by the French data protection agency (*Commission Nationale de l’Informatique et des Libertés*, CNIL).

### Statistical analysis

Statistical analyses were performed using R software (version 4.5.2; R Foundation for Statistical Computing, Vienna, Austria). Categorical variables were expressed as count (percentage, %) and continuous variables as median (interquartile range [IQR]), missing values were excluded from the denominator. Trends across ordered EHT categories were assessed using Cochran-Armitage, Jonckheere-Terpstra, or Chi-square test where appropriate. To identify factors associated with EHT level, ordinal logistic regression using the proportional-odds model was performed. Univariable analyses were first conducted, and variables with p<0·20 were entered into the multivariable model, both fitted using complete-case datasets. Odds ratios (ORs) and adjusted ORs (aORs) with 95% confidence intervals (95%CI) were reported, and the proportional-odds assumption was verified using the Brant test.

To explore the multidimensional relationships, Factor Analysis of Mixed Data (FAMD) was performed on variables with p<0·20 in univariable analysis, excluding redundant or mathematically related variables. Group separation across EHT categories was evaluated using PERMANOVA with 9,999 permutations, followed by pairwise permutational MANOVA (Pillai statistic, 9,999 permutations). p<0·05 was considered significant.

## RESULTS

### Study population and clusters

Between November 2016 and October 2025, 1,152 TB patients were diagnosed with microbiologically-confirmed TB and had available Mtb sequencing data in the eight hospitals in the Auvergne-Rhône-Alpes region, France. Consistent with regional epidemiology, 76·0% of TB patients were born outside Western Europe, most isolates belonged to lineage 4 (n=846, 73·4%) and were susceptible to first-line drugs (n=1,062, 92·2%). MDR-TB represented 41 isolates (3·6%; figure 2, table 1). For comprehensive regional MDR-TB surveillance, 25 MDR isolates diagnosed between 2010 and 2016 were included; without this addition, 16 MDR-TB cases (1·4%) would have been detected during the study period.

**Figure 2:**
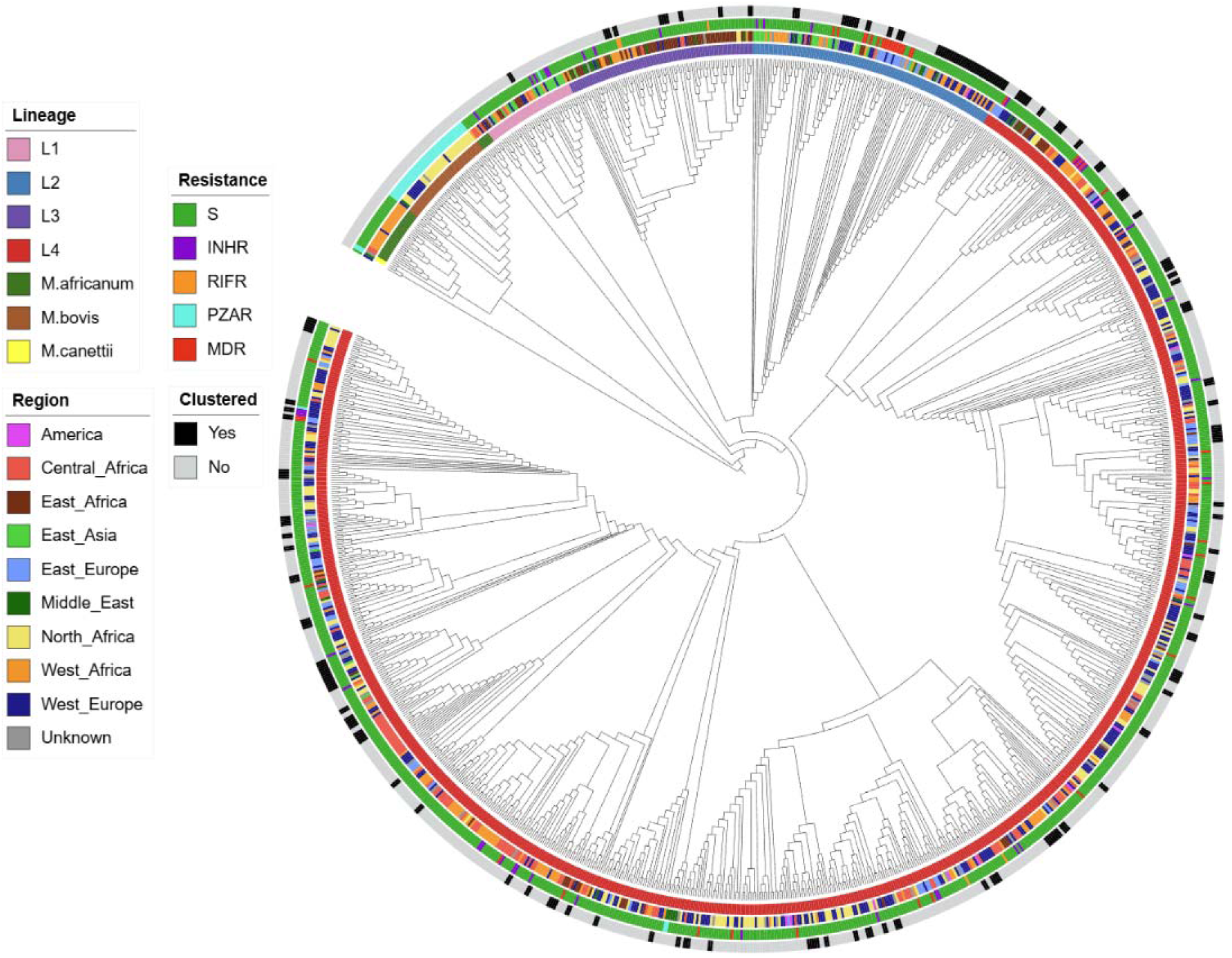
Mtb genetic diversity of the study population. Hierarchical clustering of Mtb isolates based on genomic distances. The dendrogram was generated using agglomerative hierarchical clustering with the average linkage method (hclust, R *stats* package) and subsequently converted into a phylogenetic tree using the *as.phylo* function of the *ape* package. Tree is displayed with arbitrary branch length to improve visibility. From the inside, concentric rings are coloured according to Mtb lineage, region of birth of TB patient, drug resistance profile, and genomic clustering (see legend). S: Susceptible; INHR: isoniazid (INH) mono-resistant; RIFR: rifampicin (RIF) mono-resistant; PZAR: pyrazinamide (PZA) mono-resistant; MDR: multidrug resistant meaning resistant to at least INH and RIF.

**Table 1.** Distribution of Mtb lineages and drug-resistance profiles in the study population.

|  | All isolates<br>(n=1152) | Clusters<br>(n=75) | Clustered<br>isolates<br>(n=247) | Non-clustered<br>isolates<br>(n=905) | p value<br>(clustered versus<br>non-clustered<br>isolates) |
| --- | --- | --- | --- | --- | --- |
| **Mtb complex lineage** | .. | .. | .. | .. | <0.0001 |
| Lineage 1 | 41 (3.6%) | 1 (1.3%) | 2 (0.8%) | 39 (4.3%) | 0.0058 |
| Lineage 2 | 108 (9.4%) | 6 (8.0%) | 45 (18.2%) | 63 (7.0%) | <0.0001 |
| Lineage 3 | 82 (7.1%) | 6 (8.0%) | 15 (6.1%) | 67 (7.4%) | 0.5765 |
| Lineage 4 | 846 (73.4%) | 62 (82.7%) | 185 (74.9%) | 661 (73.0%) | 0.6258 |
| *M. africanum* | 30 (2.6%) | 0 | 0 | 30 (3.3%) | 0.0011 |
| *M. bovis* | 42 (3.6%) | 0 | 0 | 42 (4.6%) | <0.0001 |
| *M. canettii* | 3 (0.3%) | 0 | 0 | 3 (0.3%) | 1.0000 |
| **Resistance profile** | .. | .. | .. | .. | 0.2435 |
| Susceptible to first-line drugs | 1062 (92.2%) | 68 (90.7%) | 232 (93.9%) | 830 (91.7%) | 0.2859 |
| INH mono-R | 38 (3.3%) | 2 (2.7%) | 4 (1.6%) | 34 (3.8%) | 0.1091 |
| RIF mono-R | 5 (0.4%) | 1 (1.3%) | 2 (0.8%) | 3 (0.3%) | 0.2923 |
| PZA mono-R (non-*M. bovis*) | 6 (0.5%) | 0 | 0 | 6 (0.7%) | 0.3511 |
| MDR | 41 (3.6%) | 4 (5.3%) | 9 (3.6%) | 32 (3.5%) | 1.0000 |

Data are presented as number of isolates (%). Clusters refer to the number of distinct genomic clusters identified, whereas clustered isolates refer to the number of isolates belonging to a genomic cluster. Comparisons between all isolates and clustered isolates were performed using the Chi-square test or Fisher's exact test, as appropriate. INH mono-R: isoniazid (INH) mono-resistant isolates; RIF mono-R: rifampicin (RIF) mono-resistant isolates; PZA mono-R (non-*M. bovis*), pyrazinamide (PZA) mono-resistant only in non-*M. bovis* isolates; MDR: multidrug-resistant tuberculosis, defined as resistance to at least INH and RIF.

All resistant isolates were detected by WGS using the 2021 WHO catalogue of mutations in Mtb complex and their association with drug resistance. The most frequent resistance-associated mutations were *inhA* c-777t and KatG S315T for isoniazid (low- or high-level resistance respectively), RpoB S450L for rifampicin, EmbB M306V for ethambutol, and PncA H57D for pyrazinamide (a polymorphism of *M. bovis* isolates). WGS also identified low-level resistance for rifampicin (due to RpoB H445N or L452P mutations) and ethambutol (EmbB M306I or Q497R mutations) undetected by the phenotypic method, as previously reported.^12^ Conversely, WGS also avoided three false resistance reports. Although the phenotypic method suggested two isoniazid_resistant and one MDR isolates, no resistance_associated mutations were detected. This discrepancy led to purity checks, revealing co_infection with non-tuberculous mycobacteria and confirming that the Mtb isolates were susceptible. Moreover, as previously reported,^13,20^ Mtb WGS also enabled the detection of resistance to a broader range of anti-TB drugs, allowing the identification of fluoroquinolone-resistant isolates (appendix pp 3-4).

Integrating genomic data with epidemiological investigations enabled the identification of 75 TB clusters (ranging from 2 to 29 patients per cluster), involving 247 TB patients, including 104 index cases and 171 secondary cases (n=28 individuals acted as both index and secondary cases). The distribution of clusters reflected the expected epidemiological success of modern Mtb lineages circulating in the region. There was no significant difference in the representation of drug_resistant isolates between clustered and non_clustered isolates (figure 2, table 1). A majority of secondary cases (120, 70·2%) were diagnosed within two years of the corresponding index case; 41 (24·0%) were household contacts, 67 (39·2%) were classified as frequent contacts and 63 (36·8%) were classified as occasional contacts (figure 3).

**Figure 3:**
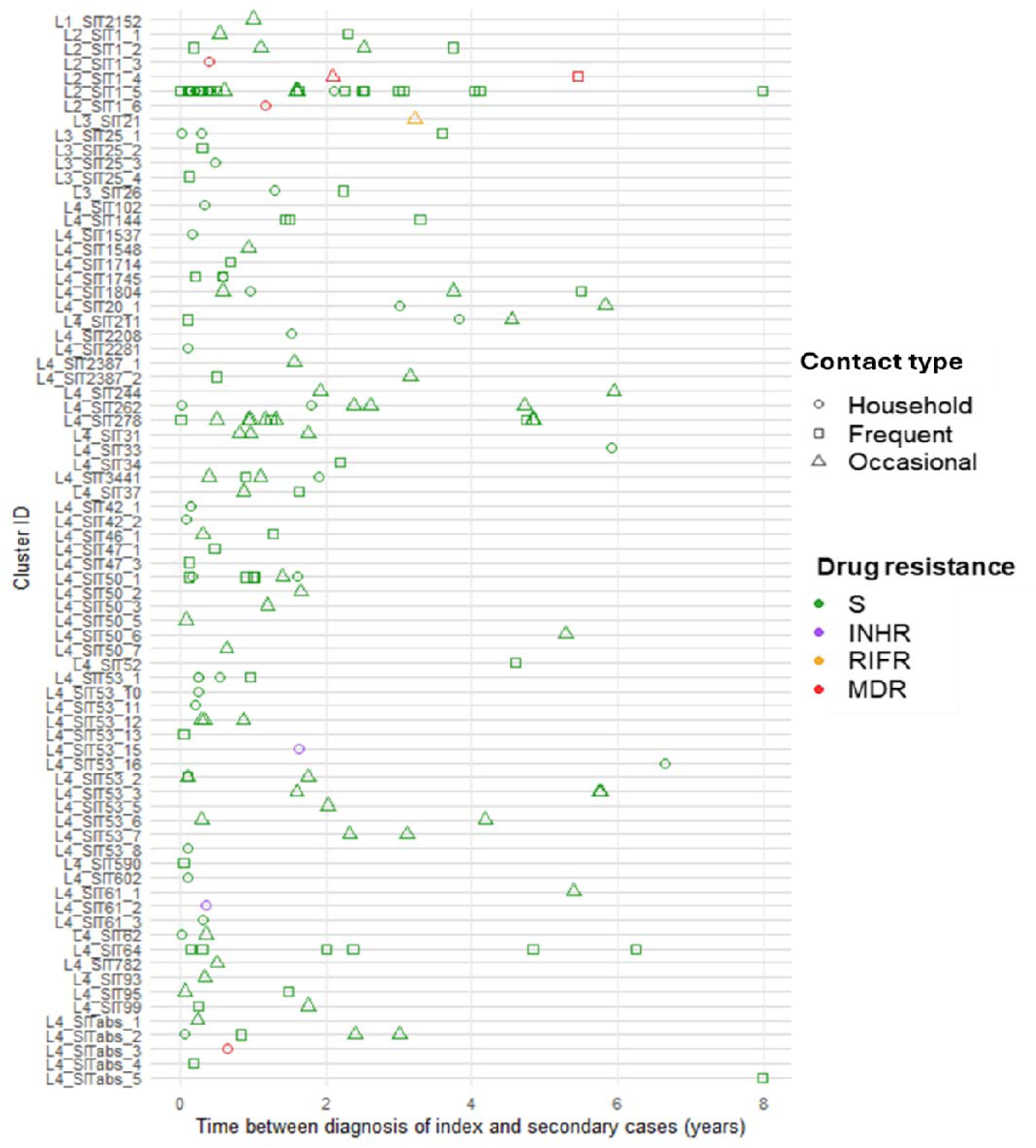
Overview of clusters. Each row represents a cluster, the cluster ID indicates Mtb lineage and the shared international type (SIT) of the cluster determined by spoligotyping. The x-axis indicates the time between diagnosis of index and secondary cases (years), each symbol represents a transmission event. Symbol forms indicate epidemiological link between index and secondary cases; circle: household contact, square: frequent contact, triangle: occasional contact. Symbol colours indicate the Mtb resistance profile. S: Susceptible; INHR: isoniazid (INH) mono-resistant; RIFR: rifampicin (RIF) mono-resistant; MDR: multidrug resistant meaning resistant to at least INH and RIF.

### Real-time impact of molecular survey

Since July 2023, the implementation of automated pairwise SNP-distance matrix generated at each WGS run has enabled real-time prospective surveillance of TB transmission. During the period from July 2023 to October 2025, 319 new TB cases were diagnosed. Among these, 54 were prospectively identified as secondary cases: 24 belonged to clusters previously recognised through retrospective analysis; 30 belonged to 23 newly identified clusters. This automated pipeline enabled clinical microbiologists to immediately interpret results, and in addition to notify TB control centres about 11 new secondary cases that led to additional field investigations (appendix p 14).

#### Nosocomial transmissions

Four prospectively detected cases corresponded to nosocomial transmissions, a setting previously investigated in our centre. In earlier work,^10^ we demonstrated that surveillance based on spoligotyping (applied to our regional database, comprising 7,563 genotyped isolates collected since 2008) enabled the identification of nosocomial transmissions, the implicated genotypes being unique within the database. This finding led to the implementation of reinforced infection-control measures to mitigate the risk of healthcare-associated TB transmissions. More recently, we reported four additional nosocomial transmission events, detected and notified thanks to the implementation of the automated pipeline.^21^ In these instances, the spoligotypes involved (Shared International Type (SIT) 1, 46, and 93, as well as one unreferenced spoligotype 700076777360771) were already represented in the database (364, 42, 26, and 2 isolates, respectively) providing no discriminatory power to identify recent transmission, except for the unreferenced spoligotype. In contrast, prospective WGS analysis provided the resolution necessary to identify these transmissions in real-time and led to prompt public-health responses.

#### Large community cluster spill over

Two of the prospectively detected cases were linked to the largest community clusters in our region. The largest is well known to the local TB control centre and has been documented in previous WGS-based investigations, including a cross-border study.^5,22^ Under the prospective system implemented, each newly detected case belonging to this cluster are automatically reported to the TB control centre. This led to extended field investigations for two patients not belonging to the main affected community, enabling assessment of potential spill over and then the identification of two additional secondary cases. Further follow-up of contacts identified during this investigation is still ongoing.

The second largest cluster also triggered notification to the TB control centre. Despite genomic evidence of ongoing transmission, these data were not incorporated into the investigation.

#### Secondary school investigation

A teenage girl was first diagnosed with TB, and initial contact investigation was restricted to her class, with no additional cases identified. Four months later, another student from a different class in the same school was diagnosed with neuro-meningeal TB. Real-time WGS demonstrated that both cases belonged to the same cluster, prompting notification to the TB control centre and extension of screening to the entire high school.

#### Reinfection versus relapse

A patient first treated for TB in 2022 later developed a new episode in 2025, but caused by a different lineage. WGS confirmed reinfection rather than relapse and identified a friend as the index case, prompting extended screening of frequent contacts of both individuals, which led to the detection of an additional case. Further follow-up of contacts identified during this investigation is still ongoing.

#### Other situations

In three additional situations, WGS enabled the identification of index cases that had not been considered during the initial investigation. First, in a retired caregiver, WGS ruled out an occupational source by linking the strain to a case diagnosed after her retirement. Second, a woman in her seventies, whose disease had been considered compatible with reactivation of childhood infection, WGS identified an index case working in a shop that the patient visited intermittently. This link could not have been established using spoligotyping alone given the high regional prevalence of SIT50 (535 isolates in the database). Finally, for a patient with miliary TB, WGS identified an index case outside the group initially screened. Both individuals were alcohol-dependent and lived within the same area and this prompted further investigations, which led to the identification of an additional TB case in the same neighbourhood.

### Index case contagiousness

While the automated pipeline enabled unbiased identification of TB transmission clusters and index cases, our objective was also to further characterise index cases with a higher level of EHT. Using the full 9-year retrospective surveillance dataset, index cases were stratified into three ordered categories reflecting increasing levels of EHT. This allowed to investigate clinical and microbiological factors associated with increasing level of EHT.

Classical indicators of infectiousness according to public health guidelines (i.e. smear microscopy, time-to-positivity, and cavitary disease),^23–25^ showed no evidence of a trend in function of EHT level (appendix pp 6-7 and 15-16). This was also the case for demographic data, substance use, co-morbidities, haematological markers of immunocompetence, as well as Mtb lineage (appendix pp 5-8). A non-significant trend (p=0.0794) between Mtb antibiotic resistance and lower EHT level was observed (appendix p 6). In contrast, the trend was significant between longer duration of symptoms before diagnosis and higher EHT level, as well as between lower severity indices in index cases (including Bandim TB score, malnutrition indicators, and C-reactive protein level) and higher EHT level (appendix pp 7-8 and 17-19). Ordinal logistic regression further supported these findings, with a trend towards increasing odds of being associated with increasing EHT level among patients presenting with prolonged TB symptoms (≥3 months, aOR=3·2, 95%CI=0·99-10·3, p=0·0529) and a significant association with clinically mild-to-moderate severity indices, such as Bandim TB score <7 (aOR=4·9, 95%CI=1·2-20·1, p=0·0285) and Malnutrition Universal Screening Tool (MUST) score <4 (aOR=16·8, 95%CI=2·9-96·3, p=0·0015, figure 4 and appendix p 9). Furthermore, FAMD using variables with the highest significance in univariable analysis (p<0·20) showed a clustering of index cases according to EHT level, with a clear separation between HH-restricted and High-EHT cases, while Low-EHT cases occupied an intermediate position (p=0·001; figure 5 and appendix p 20).

**Figure 4:**
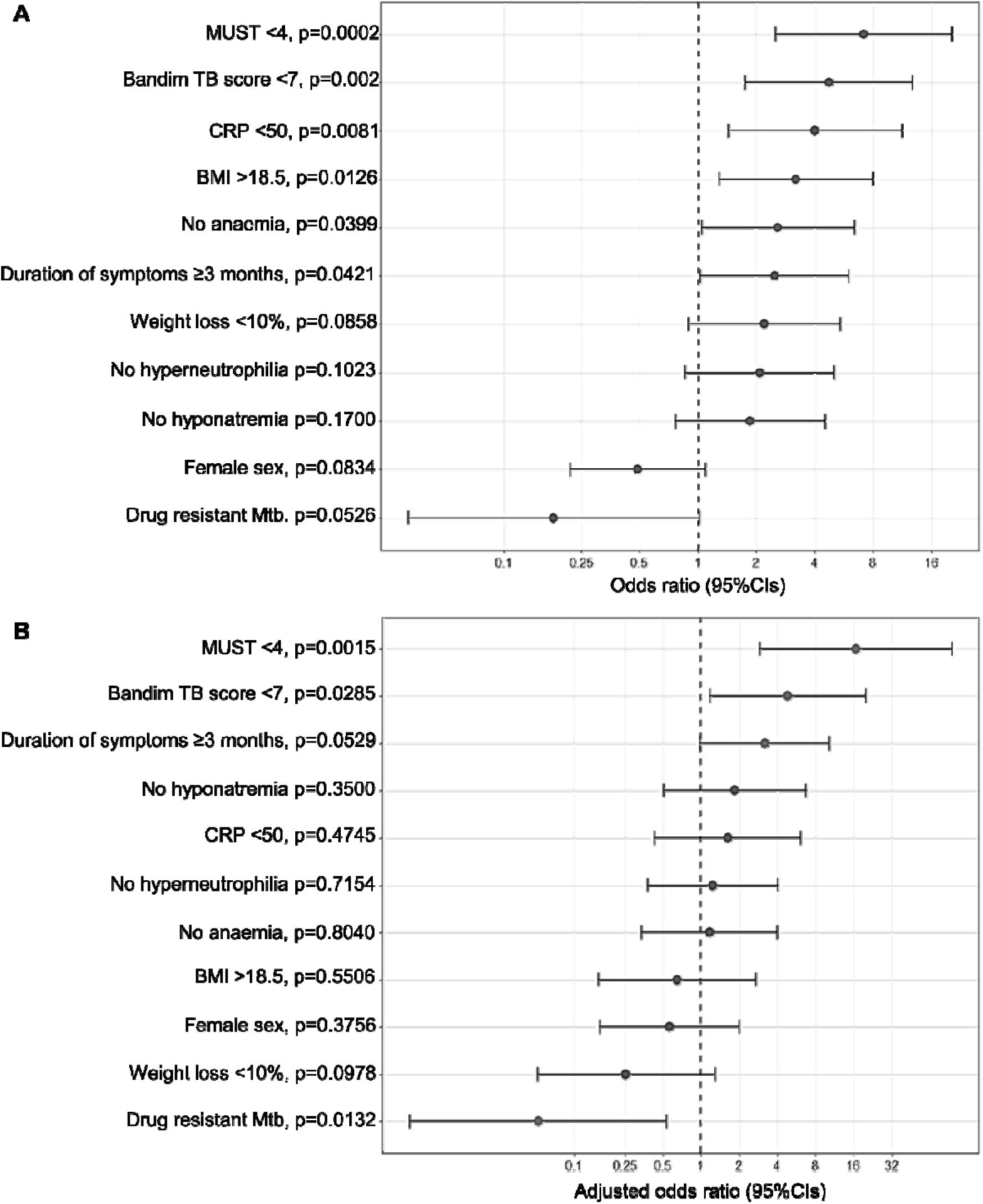
Factors associated with higher extra-household transmission (EHT) level by ordinal logistic regression. (A) Univariable proportional odds models (cumulative logit models) assessing the association between each variable and increasing level of EHT (Household-restricted [HH-restricted], Low-EHT, High-EHT). Variables included in this panel correspond to those with p<0·20 in univariable analysis. Odds ratios (ORs) with 95% confidence intervals (95%CIs) were estimated using complete-case datasets, excluding observations with missing values. The proportional odds assumption was evaluated using the Brant test. (B) Multivariable proportional odds model including all variables with p<0·20 in univariable analysis. Adjusted odds ratios (aORs) with 95% confidence intervals (95%CIs) were calculated using complete-case analysis restricted to individuals with no missing data for any included variable. Across both panels, higher ORs indicate increasing odds of being associated with increasing EHT level.

**Figure 5:**
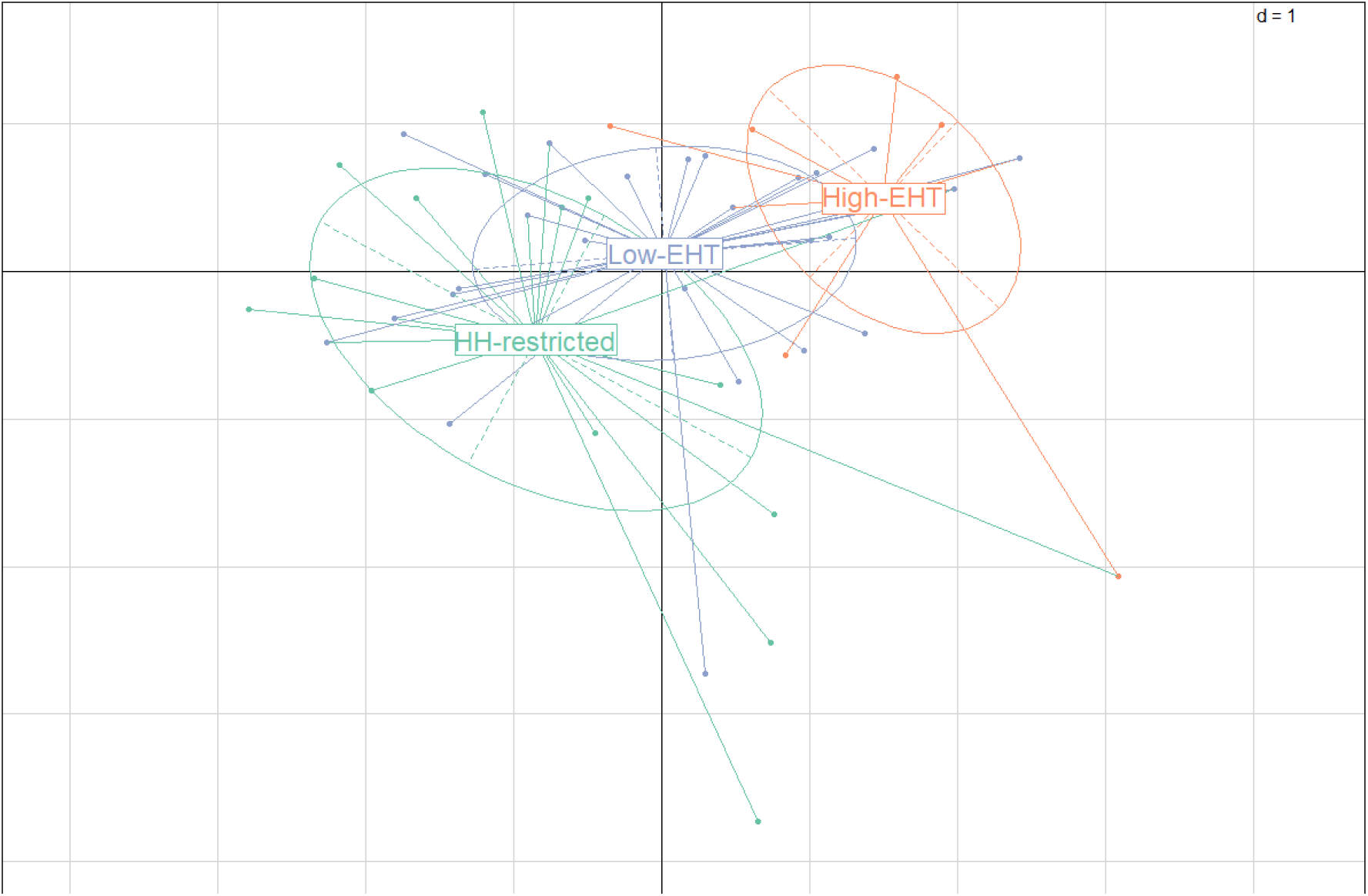
Index cases clustering according to the level of extra-household transmission (EHT) by factor analysis of mixed data (FAMD). Variables included in the analysis correspond to those with p<0·20 in univariable ordinal logistic regression, after exclusion of redundant or mathematically related variables. Observations with missing data for any included variable were excluded. Two-dimensional representation of index cases projected onto the first and third principal axes of the FAMD, explaining 29·5% and 11·9% of total variance, respectively. Ellipses represent group dispersion based on the covariance structure of each category (cellipse = 1). Group labels, HH-restricted (green), Low-EHT (blue), and High-EHT (orange), are positioned at the barycentres of their corresponding clusters. Barycentres differed significantly across groups (PERMANOVA, p=0·0001). Pairwise post-hoc comparisons confirmed significant differences between all categories (HH-restricted vs High-EHT: p=0·0003; Low-EHT vs High-EHT: p=0·0015; HH-restricted vs Low-EHT: p=0·0094).

## DISCUSSION

In this multicenter study, we report how an automated bioinformatic pipeline for anti-TB drug resistance prediction coupled with unbiased detection of TB clusters with an interface allowing direct interpretation by clinical microbiologist was implemented in routine practice in the Auvergne-Rhône-Alpes region of France. Identified TB clusters were mainly pan-susceptible lineage 4 isolates, which is consistent with epidemiology in this area.^11,26^ Among the 1152 TB cases included in this study, 21·4% were involved in a cluster and only 24·0% of transmission events were within household, corresponding to TB epidemiology in low-prevalence and high-income countries.^18,27–29^

The implementation of this pipeline allowed the identification of nosocomial transmissions that would not have been detectable through traditional field investigations. Systematic molecular epidemiological surveillance has been fundamental in enabling the triggering of investigations and thus the implementation of corrective measures in patient care.^21^ Furthermore, the integration of this pipeline in routine practice prompted the expansion of investigations through the identification of community and extra-community clusters, thus allowing the identification of epidemiological links between index and secondary cases and also the early identification of additional secondary cases. However, for one of the largest community clusters, WGS data were not incorporated into the investigation, despite genomic evidence of ongoing transmission. This likely reflects a limited consideration of genomic information in field investigations. Indeed, not all field investigators are trained to integrate WGS findings into contact tracing or to adapt their interview strategies accordingly. This gap is further reinforced by the absence of national or international recommendations on the use of WGS data for epidemiological monitoring in public health guidelines, despite the European Reference Laboratory Network for Tuberculosis (ERLTB-Net) recommending systematic WGS for TB diagnosis.^3^ Integrating WGS into public health guidelines, both for drug resistance detection and epidemiological surveillance, would provide a framework for embedding the interpretation of genomic data into the training of field investigators and for promoting its routine use in TB control activities.

Systematic surveillance also allowed the exploration of factors associated with TB transmission dynamics. Herein, we focused on contagious index TB cases, with the aim of using the level of EHT to ultimately guide field investigations towards high-risk cases. Unlike previous studies that compared clustered and non-clustered cases, or counted secondary cases without considering the links between patients or their immune status,^27,29^ this approach integrates epidemiological and clinical data to refine the assessment of transmission level. The data suggest that proxies of bacterial load do not predict the likelihood of EHT; notably, index cases with milder clinical presentations and prolonged symptoms may contribute disproportionately to EHT. This likely reflects that patients who are not malnourished and who have mild-to-moderate symptoms over an extended period maintain normal activities and social interactions,^30^ thereby exposing a broader circle of contacts to TB compared to index cases whose disease progresses rapidly to a severe form. Field interviews should therefore also consider not only the duration of symptoms but the extent to which these symptoms have affected patients’ daily activities and social behaviour. However, larger prospective studies allowing the capture of all secondary cases (active and latent TB infections [LTBI]) are needed before this is widely implemented in public health practice.

The present study is limited by its geographical scope, as it includes patients diagnosed in eight hospitals across three cities, which does not cover the entire Auvergne-Rhône-Alpes region. Notably, more than three-quarters of TB patients in the cohort were born outside Western Europe and belong to highly mobile populations, making it likely that some transmission events (especially extra-household) were missed. This high proportion of patients from middle- or high-prevalence countries also justifies our focus on active TB cases, as the elevated rates of LTBI in these groups mean that detection of LTBI status in a contact cannot be confidently attributed to recent transmission from the index case. While this strategy improves specificity, since WGS confirms that cases are caused by the same strain, it inevitably reduces sensitivity by not capturing all possible transmission events, particularly those resulting in LTBI.

Beyond its epidemiological applications, systematic WGS also enables the detection of resistance to antibiotics (a capability now considered reliable by the ERLTB-Net for first-line drugs)^3^ and accurate species identification, both of which are recommended in France in routine practice. The implementation of systematic WGS could therefore replace conventional methods for these purposes, offering a time- and cost-effective all-in-one tool.^12^ Furthermore, the approach described here provides a concrete solution for integrating Mtb WGS data into routine practice, with an automated pipeline that allows direct interpretation by clinical microbiologists, without the need for bioinformaticians at each step. In conclusion, this model could thus be extended to the national level, facilitating the widespread adoption of WGS for both epidemiological surveillance and clinical management of TB.

## Data sharing

Raw sequencing data is publicly available on the European Nucleotide Archive (https://www.ebi.ac.uk/ena/browser/home) using the accession numbers: PRJEB106242.

## Acknowledgments

This work was supported by SHAPE-Med@Lyon, a French government grant managed by the French National Research Agency under the France 2030 program (reference ANR-22-EXES-0012).

The authors thank Muriel Rabilloud (Biostatistics and Bioinformatics Department of the Lyon University Hospital) for her help in the statistical analysis of the data, Philip Robinson (DRS, Hospices Civils de Lyon, Lyon, France) for his help with manuscript preparation and the GENEPII sequencing platform (Institut des agents infectieux, Hospices Civils de Lyon, Lyon, France) for the MTBC strains sequencing.

## Contributors

CG, EH and OD were responsible for conceptualisation and methodology. The WGS data analysis pipeline was developed by CG, QT, GB-H, CBa, MV, EH and OD. Validation was performed by CG and QT. Formal analysis, and data visualisation were carried out by CG, QT, EH and OD. Investigation involved CG, CBo, OB, BJ, CT, SB, FA, CD, EH and OD. Resources were provided by CBa, OB, BJ, CT, SB, FA, CD, EH and OD. Data curation was undertaken by CG, CBo, QT, EH and OD. CG and OD wrote the original draft, and all authors reviewed and edited the manuscript.

## Supplementary Material

Supplementary appendix: Tables S1-6, Figure S1-10

