## Supplementary appendix for "Automated bioinformatic pipeline for unbiased detection of tuberculosis transmission clusters: Real-time impact and retrospective insights"

**Table of content**

### Supplementary tables

#### Table S1: Resistance mutations identified by WGS in the study population.

| **Anti-TB drug** | **Drug resistance associated mutation** | **Prevalence** |
| --- | --- | --- |
| **INH, n=79** |  |  |
|  | *inhA* c-779t | 1 (1·3%) |
|  | *inhA* c-777t | 12 (15·2%) |
|  | *inhA* c-777t + InhA I194T | 4 (5·1%) |
|  | *inhA* c-777t + InhA S94A | 3 (3·8%) |
|  | KatG DEL aa1–492 | 1 (1·3%) |
|  | KatG Q88P | 1 (1·3%) |
|  | KatG INS g2155740gc | 1 (1·3%) |
|  | KatG S315T | 48 (60·8%) |
|  | KatG S315T + fabG1 L203L | 2 (2·5%) |
|  | KatG S315T + inhA t-770a | 2 (2·5%) |
|  | KatG S315T + inhA c-777t | 3 (3·8%) |
|  | KatG R463L + ahpC t-76g | 1 (1·3%) |
| **RIF, n=46** |  |  |
|  | RpoB Q432K | 1 (2·2%) |
|  | RpoB D435V | 1 (2·2%) |
|  | RpoB D435Y | 2 (4·3%) |
|  | RpoB L443F+RpoB S450L | 1 (2·2%) |
|  | RpoB H445C | 1 (2·2%) |
|  | RpoB H445D | 3 (6·5%) |
|  | RpoB H445L | 1 (2·2%) |
|  | RpoB H445N | 1 (2·2%) * |
|  | RpoB H445Y | 2 (4·3%) |
|  | RpoB S450F | 3 (6·5%) |
|  | RpoB S450L | 28 (60·9%) |
|  | RpoB L452P | 2 (4·3%) * |
| **EMB, n=28** |  |  |
|  | EmbB N296H | 2 (7·1%) |
|  | EmbB M306V | 12 (42·9%) |
|  | EmbB M306I | 4 (14·3%) * |
|  | EmbB D354A | 4 (14·3%) |
|  | EmbB G406S | 2 (7·1%) |
|  | EmbB G406A | 1 (3·6%) |
|  | EmbB G406D | 1 (3·6%) |
|  | EmbB Q497P | 1 (3·6%) |
|  | EmbB Q497R | 1 (3·6%) * |
| **PZA, n=75** |  |  |
|  | pncA a-11g | 1 (1·3%) |
|  | pncA a-7g + PncA L19P | 1 (1·3%) |
|  | PncA L4S | 1 (1·3%) |
|  | PncA I5S + PncA T76P | 1 (1·3%) |
|  | PncA I6L | 1 (1·3%) |
|  | PncA D12A | 2 (2·7%) |
|  | PncA V21G | 1 (1·3%) |
|  | PncA A28D | 1 (1·3%) |
|  | PncA H51P | 2 (2·7%) |
|  | PncA H57D | 42 (56·0%, *M. bovis*) |
|  | PncA H57Y | 1 (1·3%) |
|  | PncA D63A | 2 (2·7%) |
|  | PncA P69S | 1 (1·3%) |
|  | PncA T87M | 1 (1·3%) |
|  | PncA K96T | 1 (1·3%) |
|  | PncA G97R | 1 (1·3%) |
|  | PncA L116P | 1 (1·3%) |
|  | PncA Q141P | 3 (4·0%) |
|  | PncA T142A | 1 (1·3%) |
|  | PncA A146T | 2 (2·7%) |
| **PZA, n=75** |  |  |
|  | PncA A146V | 1 (1·3%) |
|  | PncA T160L | 1 (1·3%) |
|  | PncA S164P | 2 (2·7%) |
|  | PncA frameshift_aa171 | 1 (1·3%) |
|  | RpsA V260I | 1 (1·3%) |
|  | PncA_WT | 2 (2·7%, *M. canettii*) |
| **FQ, n=8** |  |  |
|  | GyrA A90V | 2 (25·0%) # |
|  | GyrA A90V+GyrA D94G | 1 (12·5%) # |
|  | GyrA D94A | 1 (12·5%) $ |
|  | GyrA D94G | 2 (25·0%) #$ |
|  | GyrA D94N | 2 (25·0%) #£ |

Data were expressed as n (%). INH: isoniazid; RIF: Rifampicin; EMB: ethambutol; PZA: pyrazinamide; FQ: fluoroquinolone; *: Susceptible by phenotypic drug susceptibility testing using MGIT-960 SIRE kit, but associated with low-level resistance;^1^ #: found in multidrug-resistant (MDR) isolates; $: found in isolates susceptible to first-line anti-TB drugs; £: found in INH-resistant isolate.

#### Table S2: Demographic data of index cases

|  | **Total (n=104)** | **HH-restricted (n=31)** | **Low-EHT (n=59)** | **High-EHT (n=14)** | **p value** |
| --- | --- | --- | --- | --- | --- |
| Age | 33 (23-38) | 32 (23-38) | 33 (23-51) | 33 (20-49) | 0·4565 |
| Age class |  |  |  |  | 0·5697 |
| 0-24 | 32 (30·8%) | 10 (32·3%) | 18 (30·5%) | 4 (28·6%) |  |
| 25-44 | 37 (35·6%) | 14 (45·2%) | 18 (30·5%) | 5 (35·7%) |  |
| 45-64 | 25 (24·0%) | 4 (12·9%) | 18 (30·5%) | 3 (21·4%) |  |
| 65-95 | 10 (9·6%) | 3 (9·7%) | 5 (8·5%) | 2 (14·3%) |  |
| Sex (Male) | 70 (67·3%) | 18 (58·1%) | 40 (67·8%) | 12 (85·7%) | 0·0742 |
| Region of birth |  |  |  |  | 0·952 |
| Western Europe | 25 (24·0%) | 8 (25·8%) | 15 (25·4%) | 2 (14·3%) |  |
| Eastern Europe | 12 (11·5%) | 4 (12·9%) | 6 (10·2%) | 2 (14·3%) |  |
| North Africa | 21 (20·2%) | 6 (19·4%) | 12 (20·3%) | 3 (21·4%) |  |
| Western Africa | 17 (16·3%) | 5 (16·1%) | 8 (13·6%) | 4 (28·6%) |  |
| Central Africa | 12 (11·5%) | 3 (9·7%) | 8 (13·6%) | 1 (7·1%) |  |
| Eastern Africa | 14 (13·5%) | 5 (16·1%) | 8 (13·6%) | 1 (7·1%) |  |
| Middle East Asia | 3 (2·9%) | 0 (0%) | 2 (3·4%) | 1 (7·1%) |  |

Data were expressed as n (%) or median (IQR). Index cases were stratified into three ordered extra‑household transmission (EHT) categories: household-restricted (HH-restricted), low-EHT, high-EHT. Trends across the ordered EHT categories were assessed using the Jonckheere–Terpstra or Chi‑square test, as appropriate.

#### Table S3: Microbiological data of index cases

|  | **Total (n=104)** | **HH-restricted (n=31)** | **Low-EHT (n=59)** | **High-EHT (n=14)** | **p value** |
| --- | --- | --- | --- | --- | --- |
| Lineage |  |  |  |  | 0·5362 |
| L1 | 1 (1·0%) | 0 (0%) | 1 (1·7%) | 0 (0%) |  |
| L2 | 14 (13·5%) | 6 (19·4%) | 5 (8·5%) | 3 (21·4%) |  |
| L3 | 8 (7·7%) | 3 (9·7%) | 5 (8·5%) | 0 (0%) |  |
| L4 | 81 (77·9%) | 22 (71·0%) | 48 (81·3%) | 11 (78·6%) |  |
| Resistance |  |  |  |  | 0·0794 |
| Susceptible | 97 (93·3%) | 26 (83·9%) | 58 (98·3%) | 13 (92·9%) |  |
| INH-Resistant | 2 (1·9%) | 2 (6·5%) | 0 (0%) | 0 (0%) |  |
| RIF-Resistant | 1 (1·0%) | 0 (0%) | 1 (1·7%) | 0 (0%) |  |
| MDR | 4 (3·8%) | 3 (9·7%) | 0 (0%) | 1 (7·1%) |  |
| α-diversity >1 | 41 (39·4%) | 10 (32·3%) | 25 (42·4%) | 6 (42·9%) | 0·3948 |
| Positive smear result | 68/96 (70·8%) | 21/29 (72·4%) | 39/54 (72·2%) | 8/13 (61·5%) | 0·5588 |
| Time-to-positivity (days) | 6 (4-10) | 5·5 (4-9·5) | 6 (4·5-9) | 8 (4-11) | 0·4292 |

Data were expressed as n (%), n/N (%) when missing data were present, or median (IQR). Missing data were excluded from denominators. Index cases were stratified into three ordered extra‑household transmission (EHT) categories: household-restricted (HH-restricted), low EHT, high EHT. Trends across the ordered EHT categories were assessed using the Cochran-Armitage, Jonckheere-Terpstra, or Chi‑square test, as appropriate. Susceptible: susceptible to first-line drugs; INH-Resistant, isoniazid (INH) mono-resistant; RIF-Resistant: rifampicin (RIF) mono-resistant; MDR, multidrug-resistant meaning resistant to at least INH and RIF; α-diversity >1: Mtb isolates for which micro-diversity was detected, as previously described.^2^

#### Table S4: TB clinical presentation, comorbidities and substance use of index cases

|  | **Total** | **HH-restricted** | **Low-EHT** | **High-EHT** | **p value** |
| --- | --- | --- | --- | --- | --- |
| **Clinical presentation** |  |  |  |  |  |
| Cavitary disease | 65/85 (76·5%) | 20/25 (80·0%) | 35/47 (74·5%) | 10/13 (76·9%) | 0·7473 |
| Extrapulmonary TB | 15/79 (19·0%) | 4/23 (17·4%) | 7/43 (16·3%) | 4/13 (30·8%) | 0·4114 |
| Duration of symptoms before diagnosis (month) | 2 (1-4) | 1·5 (1-4) | 2·5 (1-4) | 4 (1·5-6) | 0·0637 |
| Duration of symptoms ≥3 months | 37/79 (46·8%) | 8/24 (33·3%) | 20/42 (47·6%) | 9/13 (69·2%) | 0·0384 |
| Bandim TB score | 5 (4-7) | 7 (4-8) | 5 (5-7) | 4 (2·5-5) | 0·0007 |
| Bandim TB score class |  |  |  |  | 0·0050 |
| Paucisymptomatic (≤2) | 8/75 (10·7%) | 2/23 (8·7%) | 3/39 (7·7%) | 3/13 (23·1%) |  |
| Mild (3-4) | 17/75 (22·7%) | 5/23 (21·7%) | 6/39 (15·4%) | 6/13 (46·2%) |  |
| Moderate (5-6) | 26/75 (34·7%) | 4/23 (17·4%) | 18/39 (46·2%) | 4/13 (30·8%) |  |
| Severe (≥7) | 24/75 (32·0%) | 12/23 (52·2%) | 12/39 (30·8%) | 0/13 (0%) |  |
| CRP (mg/L) | 85 (39-134) | 97 (81-154) | 84 (30-119) | 42 (21-107) | 0·0046 |
| CRP ≥50mg/L | 49/72 (68·1%) | 19/22 (86·4%) | 25/38 (65·8%) | 5/12 (41·7%) | 0·0069 |
| **Comorbidities** |  |  |  |  |  |
| HIV | 1/77 (1·3%) | 0/23 (0%) | 1/41 (2·4%) | 0/13 (0%) | 0·8456 |
| Hepatitis | 2/77 (2·6%) | 1/23 (4·3%) | 1/41 (2·4%) | 0/13 (0%) | 0·4295 |
| Diabetes | 8/77 (10·4%) | 2/23 (8·7%) | 4/41 (9·8%) | 2/13 (15·4%) | 0·5322 |
| Immunosuppressive treatment | 1/77 (1·3%) | 1/23 (4·3%) | 0/41 (0%) | 0/13 (0%) | 0·1920 |
| History of TB | 9/77 (11·7%) | 2/23 (8·7%) | 4/41 (9·8%) | 3/13 (23·1%) | 0·2518 |
| **Substance use** |  |  |  |  |  |
| Tobacco | 41/78 (52·6%) | 12/23 (52·2%) | 22/42 (52·4%) | 7/13 (53·8%) | 0·9306 |
| Alcohol | 17/78 (21·8%) | 4/23 (17·4%) | 9/42 (21·4%) | 4/13 (30·8%) | 0·3703 |
| Drugs | 3/78 (3·8%) | 3/23 (13·0%) | 0/42 (0%) | 0/13 (0%) | 0·0210 |

Data were expressed as n/N (%) or median (IQR). Missing data were excluded from denominators. Index cases were stratified into three ordered extra‑household transmission (EHT) categories: household-restricted (HH-restricted), low EHT, high EHT. Trends across the ordered EHT categories were assessed using the Cochran-Armitage, Jonckheere-Terpstra, or Chi‑square test, as appropriate. The Bandim TB score considers five symptoms (cough, haemoptysis, dyspnoea, chest pain, night sweats) and five clinical findings (anaemia, tachycardia, positive finding at lung auscultation, fever, body mass index (BMI) <18 and <16); one point is attributed for each aspect, and the final score is the sum of these. CRP: C-reactive protein; HIV: Human immunodeficiency virus coinfection; Hepatitis: active hepatitis B or C coinfection; Drugs: intravenous or inhaled drug users.

#### Table S5: Haematological markers and nutritional status of index cases

|  | **Total (n=75)** | **HH-restricted (n=22)** | **Low-EHT (n=40)** | **High-EHT (n=13)** | **p value** |
| --- | --- | --- | --- | --- | --- |
| **Haematological markers** |  |  |  |  |  |
| Haemoglobin (g/L) | 116 (99-131) | 97 (84-120) | 120 (101-136) | 121 (114-130) | 0·0025 |
| Anaemia | 43/75 (57·3%) | 17/22 (73·9%) | 20/40 (50·0%) | 6/13 (46·2%) | 0·0426 |
| Leukocytes (G/L) | 8·8 (6·5-10·9) | 9·9 (6·3-12·2) | 7·8 (6·6-10·6) | 9·1 (6·5-10·7) | 0·6850 |
| Hyperleukocytosis (>11G/L) | 17/75 (22·7%) | 7/22 (31·8%) | 8/40 (20·0%) | 2/13 (15·4%) | 0·2248 |
| Neutrophils (G/L) | 6·2 (4·6-8·6) | 7·1 (4·5-9·3) | 6·1 (4·3-8·4) | 5·7 (4·6-7·7) | 0·3943 |
| Hyperneutrophilia (>7.5G/L) | 33/75 (44·0%) | 13/22 (59·1%) | 16/40 (40·0%) | 4/13 (30·8%) | 0·1058 |
| Monocytes (G/L) | 0·82 (0·61-1·02) | 0·83 (0·49-1·03) | 0·80 (0·61-1·02) | 0·81 (0·64-1·12) | 0·6887 |
| Hypermonocytosis (>1G/L) | 21/75 (28·0%) | 6/22 (27·3%) | 11/40 (27·5%) | 4/13 (30·8%) | 0·8424 |
| Lymphocytes (G/L) | 1·3 (0·97-1·9) | 1·4 (0·99-2·0) | 1·3 (0·97-1·9) | 1·5 (1·0-2·0) | 0·8631 |
| Lymphopenia (<1G/L) | 21/75 (28·0%) | 6/22 (27·3%) | 12/40 (30%) | 3/13 (23·1%) | 0·8544 |
| NLR | 4·1 (3·0-8·1) | 4·0 (3·5-11·3) | 3·9 (3·0-7·8) | 4·8 (2·8-5·1) | 0·3509 |
| NLR ≥5 | 31/75 (41·3%) | 10/22 (45·5%) | 18/40 (45·0%) | 3/13 (23·1%) | 0·2528 |
| LMR | 1·8 (1·0-2·4) | 2·1 (1·0-2·4) | 1·7 (1·0-2·4) | 1·7 (1·3-3·0) | 0·9031 |
| LMR ≤1·5 | 33/75 (44·0%) | 7/22 (31·8%) | 20/40 (50·0%) | 6/13 (46·2%) | 0·3059 |
| **Malnutrition markers** |  |  |  |  |  |
| BMI kg/m² | 19·1 (17·6-21·0) | 18·3 (16·7-19·7) | 19·0 (17·4-20·42) | 21·6 (20·3-22·6) | 0·0021 |
| BMI ≤18·5 | 33/75 (44·0%) | 13/22 (59·1%) | 19/40 (47·5%) | 1/13 (7·7%) | 0·0087 |
| Unintentional weight loss (%) | 10·7 (5·6-19·2) | 16·0 (7·0-20·4) | 10·5 (7·3-20·0) | 5·9 (3·8-13·7) | 0·0376 |
| Weight loss ≤10% | 45/75 (60%) | 16/22 (72·3%) | 24/40 (60·0%) | 5/13 (38·5%) | 0·0793 |
| MUST | 4 (2-6) | 5·5 (4-6) | 4 (2-5) | 1 (1-2) | <0·0001 |
| MUST ≥4 | 41/75 (54·7%) | 17/22 (77·3%) | 24/40 (60·0%) | 0/13 (0%) | 0·0001 |

Data were expressed as n/N (%) or median (IQR). Missing data were excluded from denominators. Index cases were stratified into three ordered extra‑household transmission (EHT) categories: household-restricted (HH-restricted), low EHT, high EHT. Trends across the ordered EHT categories were assessed using the Cochran-Armitage, Jonckheere-Terpstra test, or Chi‑square test, as appropriate. NLR: Neutrophil to Lymphocyte Ratio; LMR: Lymphocyte to Monocyte Ratio; BMI: Body Mass Index: MUST: Malnutrition Universal Screening Tool that includes three variables, unintentional weight loss score (weight loss <5% = 0, weight loss 5-10% = 1, weight loss >10% = 2), BMI (>20·0 = 0·18·5-20·0 = 1, <18·5 = 2), and anorexia (if yes = 2), and the final score is the sum of these.

#### Table S6: Ordinal logistic regression analysis of factors associated with extra-household transmission (EHT) level

|  | **OR (95%CI)** | **p value** | **aOR (95%CI)** | **p value** |
| --- | --- | --- | --- | --- |
| **Demographic data** |  |  |  |  |
| Age class | 0·8 (0·3-2·1) | 0·6739 | ·· | ·· |
| Sex (F) | 0·5 (0·2-1·1) | 0·0834 | 0·6 (0·2-2·0) | 0·3756 |
| Region of birth | 1·3 (0·3-4·7) | 0·7272 | ·· | ·· |
| **Microbiological data** |  |  |  |  |
| Lineage | 1·5 (0·05-48·7) | 0·8171 | ·· | ·· |
| Drug-resistant Mtb | 0·18 (0·03-1·0) | 0·0526 | 0·05 (0·01-0·54) | 0·0132 |
| α-diversity >1 | 0·7 (0·3-1·5) | 0·3780 | ·· | ·· |
| Smear positive | 0·8 (0·3-2·0) | 0·5851 | ·· | ·· |
| Time to positivity <7 days | 0·8 (0·4-2·1) | 0·7271 | ·· | ·· |
| **Clinical presentation** |  |  |  |  |
| Cavitary disease | 0·8 (0·3-2·6) | 0·7292 | ·· | ·· |
| Extrapulmonary TB | 1·6 (0·4-4·8) | 0·4305 | ·· | ·· |
| Duration of symptoms ≥3 months | 2·5 (1·0-6·0) | 0·0421 | 3·2 (0·99-10·3) | 0·0529 |
| Bandim TB score <7 | 4·8 (1·8-12·8) | 0·002 | 4·9 (1·2-20·1) | 0·0285 |
| CRP <50mg/L | 4·0 (1·4-11·3) | 0·0081 | 1·6 (0·4-6·2) | 0·4745 |
| No hyponatremia | 1·9 (0·8-4·5) | 0·17 | 1·9 (0·5-6·7) | 0·35 |
| No hypochloraemia | 1·4 (0·5-4·0) | 0·4829 | ·· | ·· |
| **Comorbidities** |  |  |  |  |
| Hepatitis | 2·8 (0·2-39·6) | 0·4503 | ·· | ·· |
| Diabetes | 0·6 (0·2-2·7) | 0·5402 | ·· | ·· |
| History of TB | 0·4 (0·1-1·8) | 0·2568 | ·· | ·· |
| **Substance use** |  |  |  |  |
| Tobacco | 1·0 (0·4-2·3) | 0·9333 | ·· | ·· |
| Alcohol | 1·7 (0·6-5·0) | 0·3772 | ·· | ·· |
| **Haematological markers** |  |  |  |  |
| No anaemia | 2·6 (1·0-6·5) | 0·0399 | 1·2 (0·3-4·0) | 0·804 |
| Leukocytes <11G/L | 1·9 (0·7-5·5) | 0·2161 |  |  |
| Neutrophils <7.5G/L | 2·1 (0·9-5·1) | 0·1023 | 1·2 (0·4-4·1) | 0·7154 |
| Monocytes <1G/L | 0·9 (0·3-2·4) | 0·8495 | ·· | ·· |
| Lymphocytes >1G/L | 1·1 (0·4-2·8) | 0·8750 | ·· | ·· |
| NLR <5 | 1·6 (0·7-3·9) | 0·2794 | ·· | ·· |
| LMR >1·5 | 0·6 (0·3-1·5) | 0·2848 | ·· | ·· |
| **Nutritional status** |  |  |  |  |
| BMI >18·5kg/m² | 3·2 (1·3-8·0) | 0·0126 | 0·6 (0·2-2·7) | 0·5506 |
| Unintentional weight loss <10% | 2·2 (0·9-5·4) | 0·0858 | 0·3 (0·1-1·3) | 0·0978 |
| MUST <4 | 7·2 (2·5-20·4) | 0·0002 | 16·8 (2·9-96·3) | 0·0015 |

Index cases were stratified into three ordered extra‑household transmission (EHT) categories: household-restricted (HH-restricted), low EHT, high EHT. To identify factors associated with EHT level, ordinal logistic regression using the proportional odds model was performed. Univariable analyses were first conducted, and variables with p<0·20 were entered into the multivariable model, both fitted using complete-case datasets. Odds ratios (ORs) and adjusted odds ratios (aORs) with 95% confidence intervals (95%CI) were reported. The proportional odds assumption was assessed using the Brant test. Drug resistant Mtb: INH mono-resistant or RIF mono-resistant or MDR.

### Supplementary figures

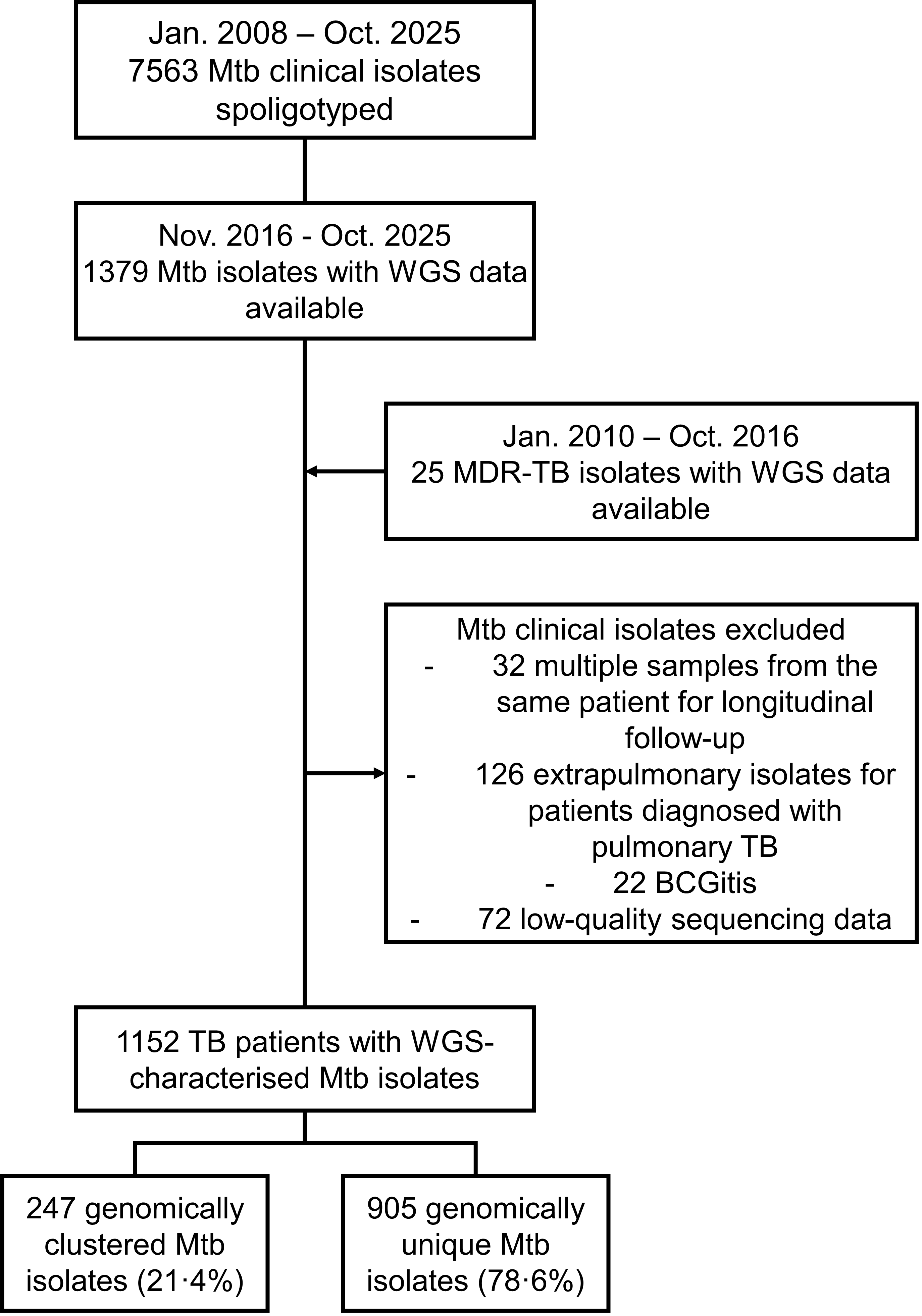

#### Figure S1: Flow diagram of sample inclusion.

MDR-TB: Multidrug-resistant tuberculosis, meaning resistant to at least rifampicin and isoniazid; Mtb: *Mycobacterium tuberculosis*; TB: tuberculosis; WGS: Whole-genome sequencing.

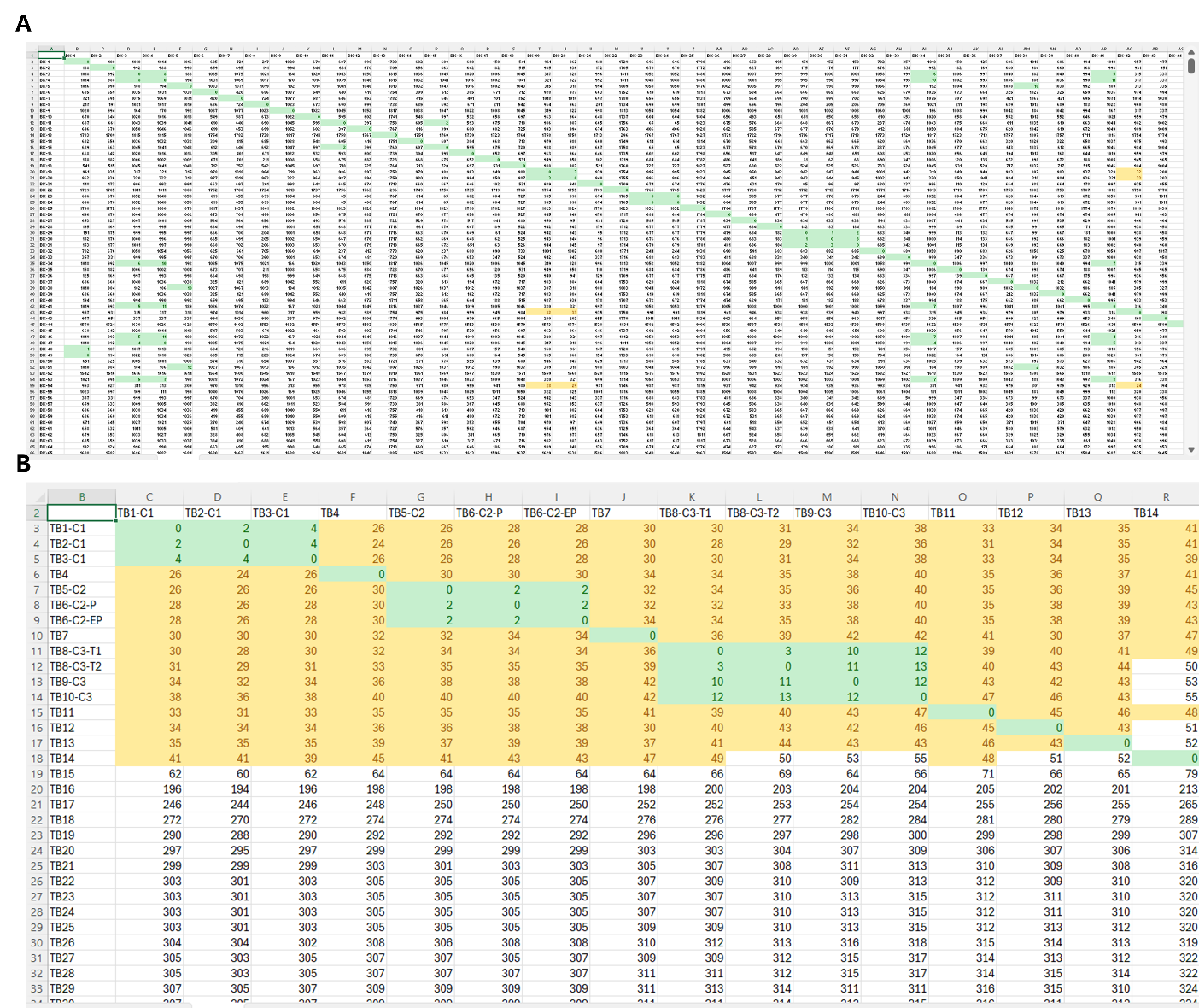

#### Figure S2: Distance matrix highlighting clades and transmission clusters based on pairwise SNP distances.

(A) Overview of pairwise single nucleotide polymorphism (SNP) distance matrix between clinical *Mycobacterium tuberculosis* (Mtb) isolates. (B) Zoomed view of the distance matrix focusing on samples belonging to clusters (pairwise SNP distance ≤20, highlight in green) and/or clades (pairwise SNP distance between 21 and 50, highlighted in yellow). Samples highlighted in green were selected for a manual curation step based on alignment inspection using the Integrative Genome Viewer (IGV)-report to validate SNP distances.

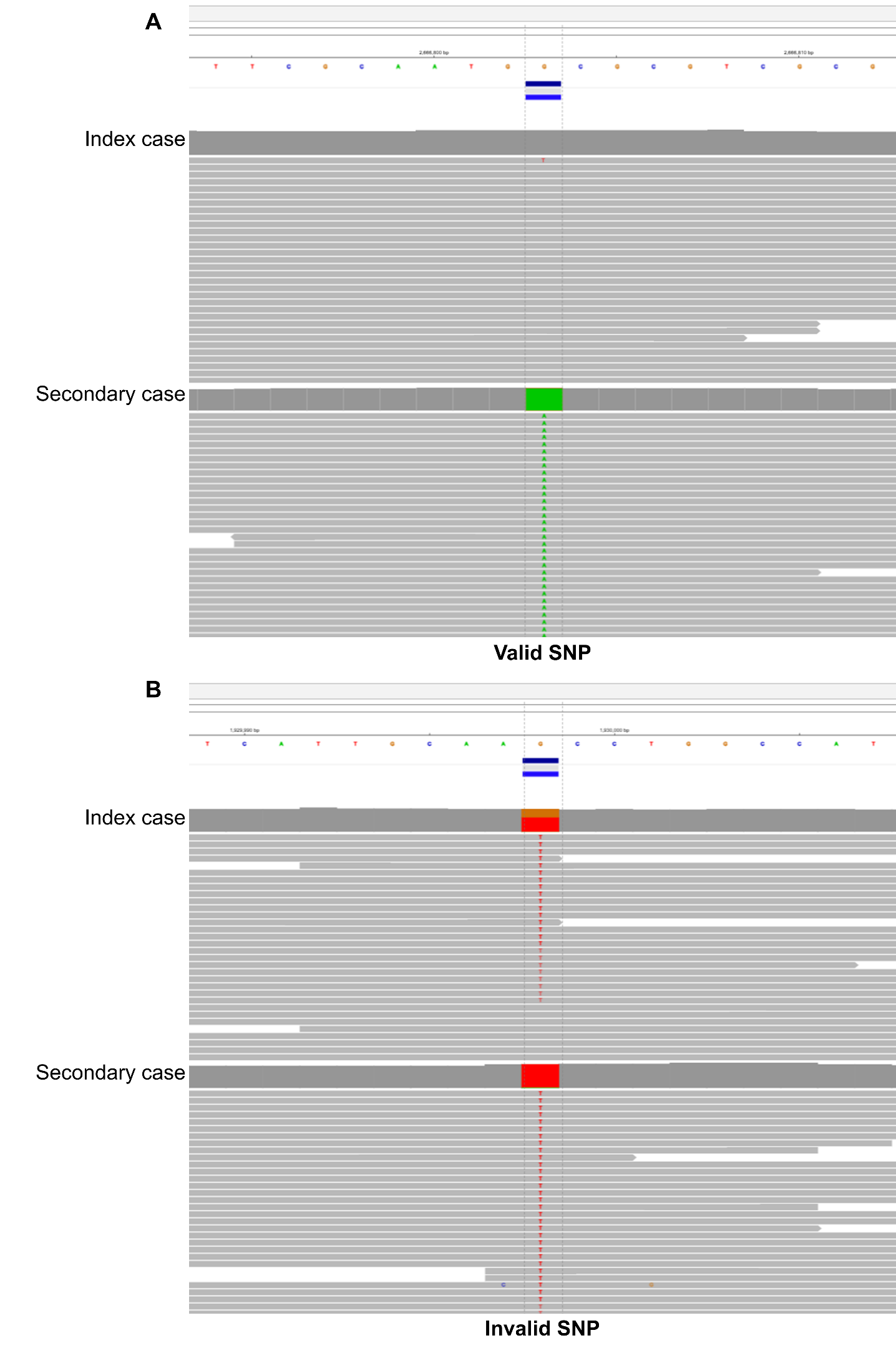

#### Figure S3: Integrative Genome Viewer (IGV) alignments illustrating validation of pairwise SNP distances between index and secondary cases.

Representative screenshots of read alignments visualised in IGV showing pairwise SNP distance assessment between putative index and secondary TB cases. (A) Example of a valid pairwise SNP distance, with a fixed mutation present in the secondary case isolate and absent in the index case isolate. (B) Example of an invalid pairwise SNP distance, where the mutation is detected in the secondary case but did not pass variant calling filters in the index case because it unfixed. The within-host Mtb micro-diversity passed from the index to the secondary case was excluded from pairwise SNP distance analyses and was instead leveraged as an indicator to refine reconstruction of transmission links.

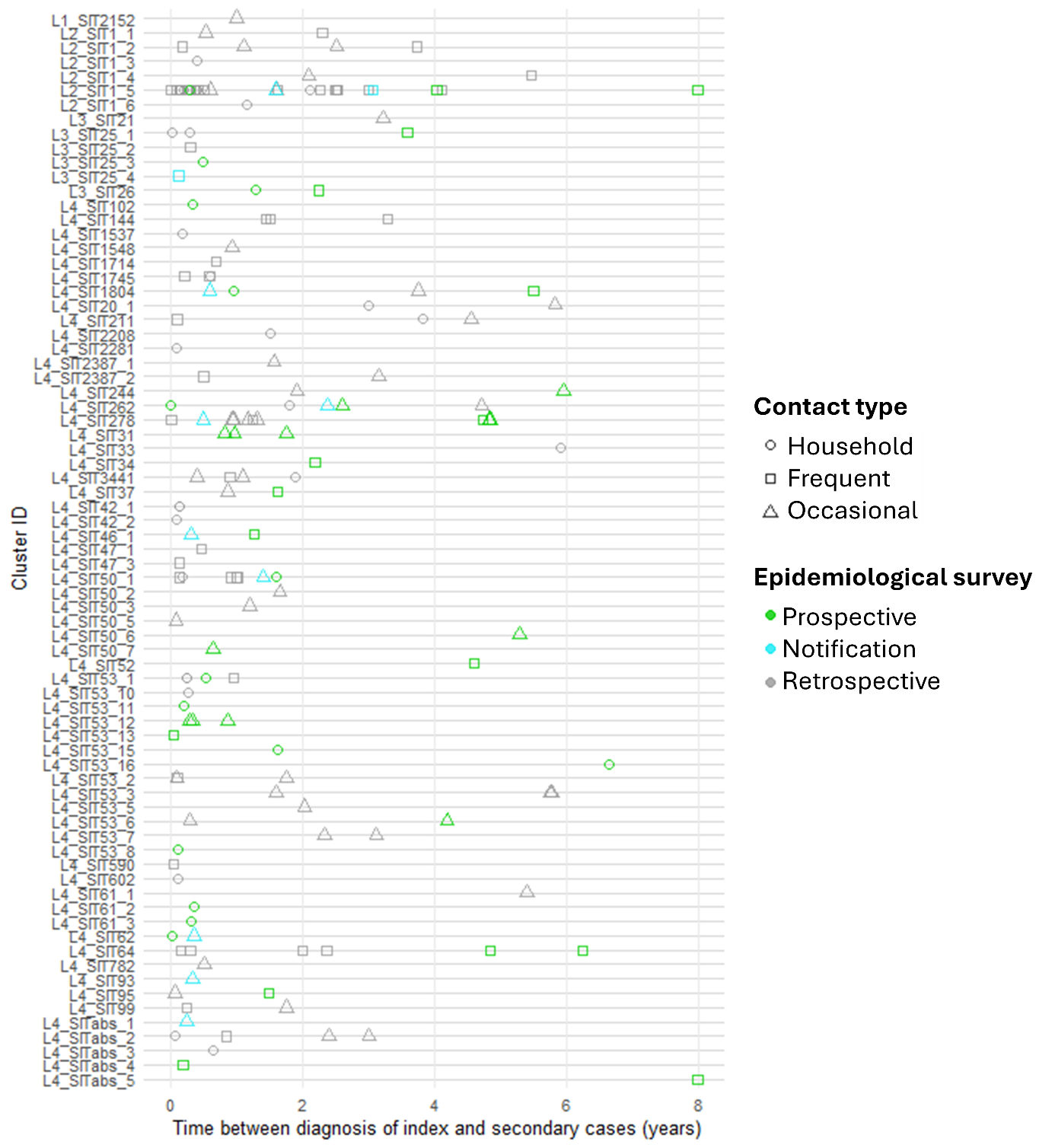

#### Figure S4: Retrospective and prospective identification of clustered cases.

Each row represents a cluster with cluster ID indicating Mtb lineage and the shared international type (SIT) of the cluster determined by spoligotyping. The x-axis indicates the time between diagnosis of index and secondary cases (years), each symbol representing a transmission event. Symbol forms indicate epidemiological link between index and secondary cases; circle: household contact, square: frequent contact, triangle: occasional contact. Symbol colours indicate whether transmission event was identified during retrospective (grey) or prospective (green or cyan) surveillance, and whether its identification led to notification to field investigators (cyan).

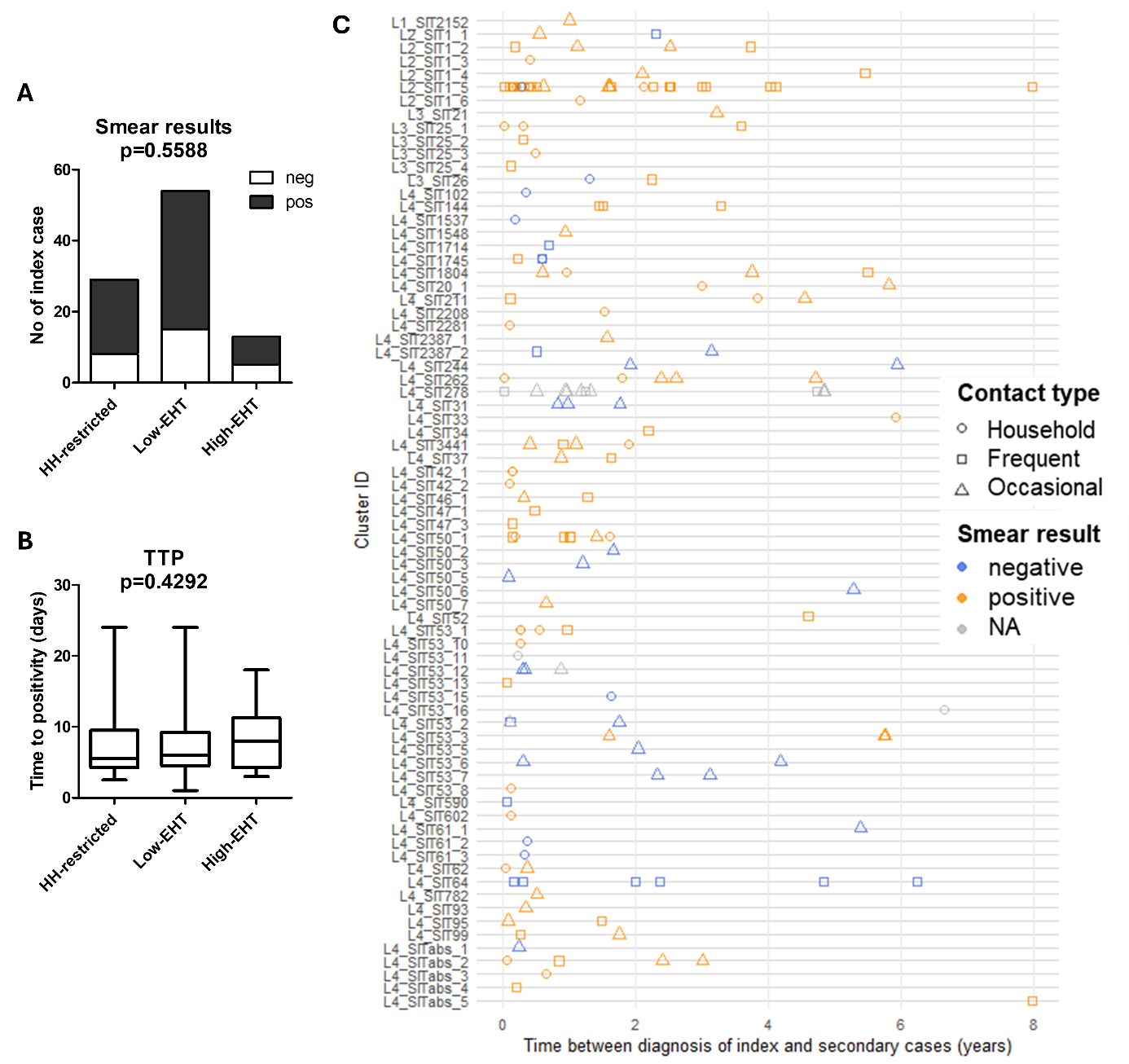

#### **Figure S5: Trend in bacterial load in clinical samples of index cases according to EHT level.**

Index cases were stratified into three ordered categories according to their level of extra household transmission (EHT), by considering those transmitting TB: exclusively within household (HH-restricted), to at least one immunocompetent secondary case outside of household (Low-EHT) and to two or more immunocompetent secondary cases outside of household (High-EHT). (A) Trend in smear microscopy results according to EHT level, assessed using the Cochran‑Armitage test. (B) Trend in the median time to positivity of liquid cultured clinical samples according to EHT level, assessed using the Jonckheere-Terpstra test. (C) Each row represents a cluster with cluster ID indicating Mtb lineage and the shared international type (SIT) of the cluster determined by spoligotyping. The x-axis indicates the time between diagnosis of index and secondary cases (years), each symbol representing a transmission event. Symbol forms indicate epidemiological link between index and secondary cases; circle: household contact, square: frequent contact, triangle: occasional contact. Symbol colours indicate the smear status of the index case: positive (orange), negative (blue), or not available data (NA; grey).

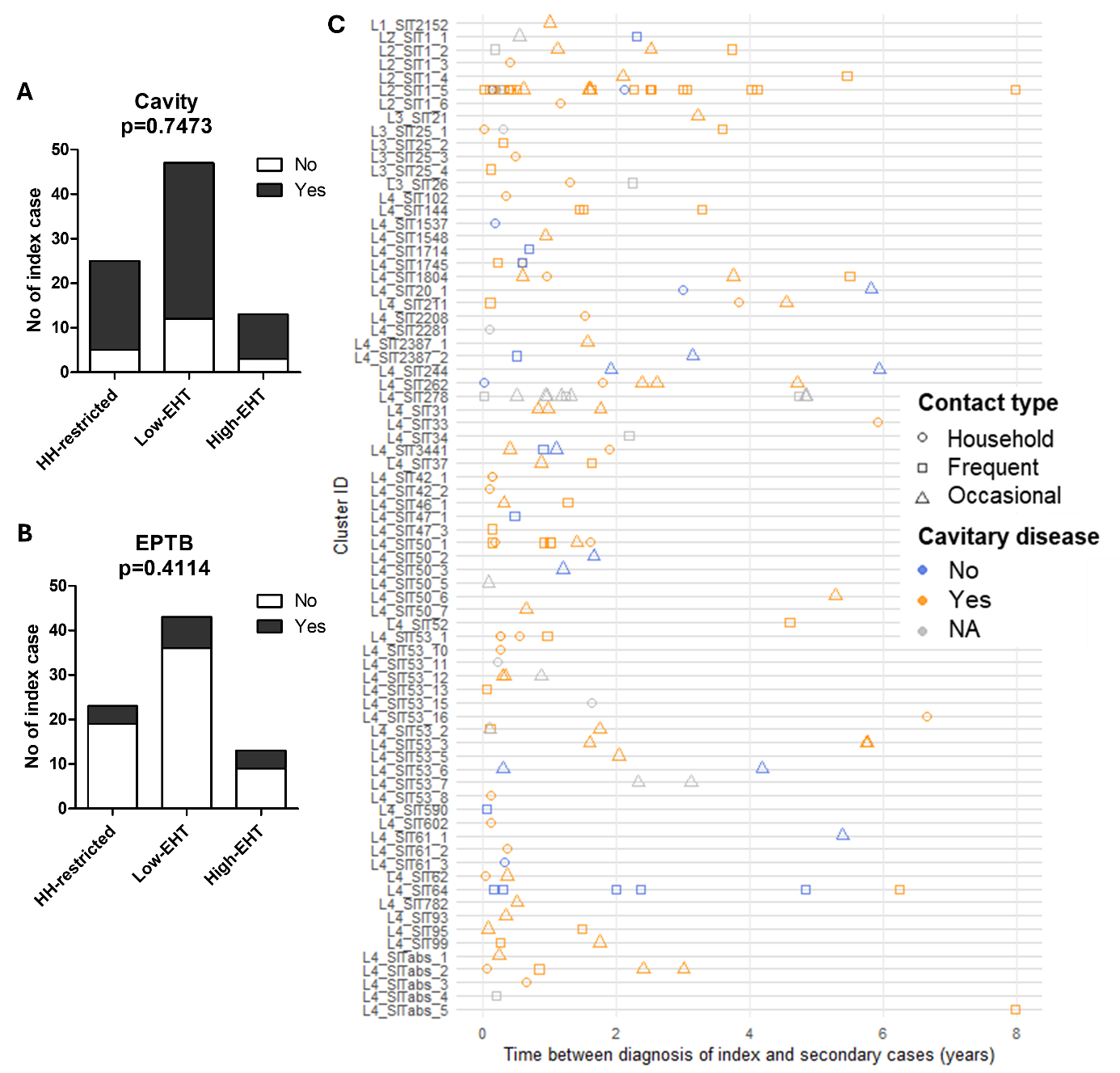

#### **Figure S6: Trend in TB clinical presentation in index cases according to EHT level.**

Index cases were stratified into three ordered categories according to their level of extra household transmission (EHT), by considering those transmitting TB: exclusively within household (HH-restricted), to at least one immunocompetent secondary case outside of household (Low-EHT) and to two or more immunocompetent secondary cases outside of household (High-EHT). (A) Trend in cavitary disease in index cases according to EHT level. (B) Trend in extrapulmonary TB (EPTB) dissemination in index cases according to EHT level. (A-B) Trends across ordered EHT categories were assessed using the Cochran-Armitage test. (C) Each row represents a cluster with cluster ID indicating Mtb lineage and the shared international type (SIT) of the cluster determined by spoligotyping. The x-axis indicates the time between diagnosis of index and secondary cases (years), each symbol representing a transmission event. Symbol forms indicate epidemiological link between index and secondary cases; circle: household contact, square: frequent contact, triangle: occasional contact. Symbol colours indicate the cavitary status of the index case: Yes (orange), No (blue), or not available data (NA; grey).

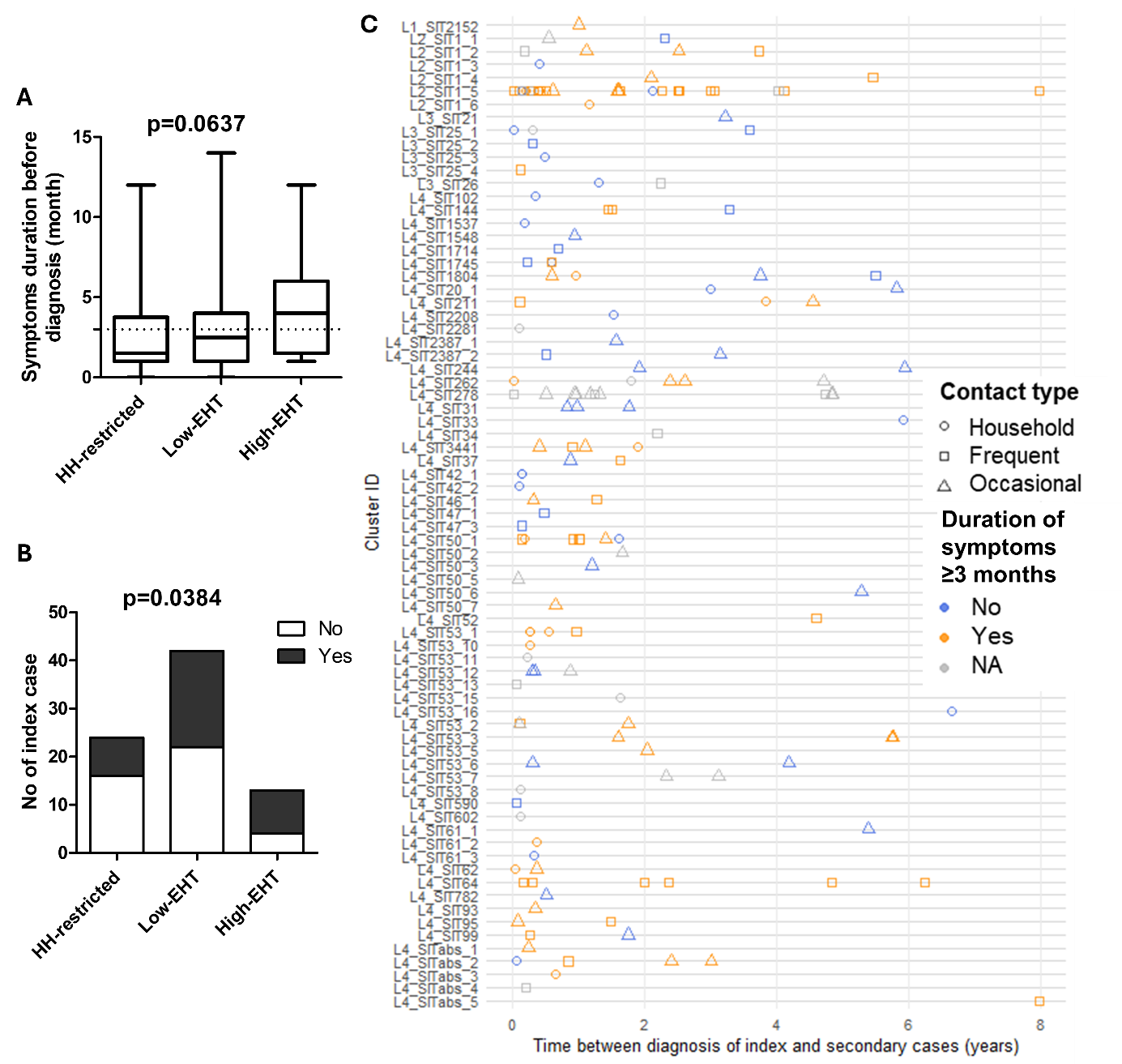

Figure S7: Trend in duration of symptoms before diagnosis of index cases according to EHT level.

Index cases were stratified into three ordered categories according to their level of extra household transmission (EHT), by considering those transmitting TB: exclusively within household (HH-restricted), to at least one immunocompetent secondary case outside of household (Low-EHT) and to two or more immunocompetent secondary cases outside of household (High-EHT). (A) Trend in the median duration of symptoms before diagnosis in index cases according to EHT level, assessed using the Jonckheere-Terpstra test. (B) Trend in duration of symptoms before diagnosis of index cases ≥3 months (Yes: black; No: white) according to EHT level, assessed using the Cochran-Armitage test. (C) Each row represents a cluster with cluster ID indicating Mtb lineage and the shared international type (SIT) of the cluster determined by spoligotyping. The x-axis indicates the time between diagnosis of index and secondary cases (years), each symbol representing a transmission event. Symbol forms indicate epidemiological link between index and secondary cases; circle: household contact, square: frequent contact, triangle: occasional contact. Symbol colours indicate duration of symptoms before diagnosis of the index case: ≥3 months (orange), <3 months (blue), or not available data (NA; grey).

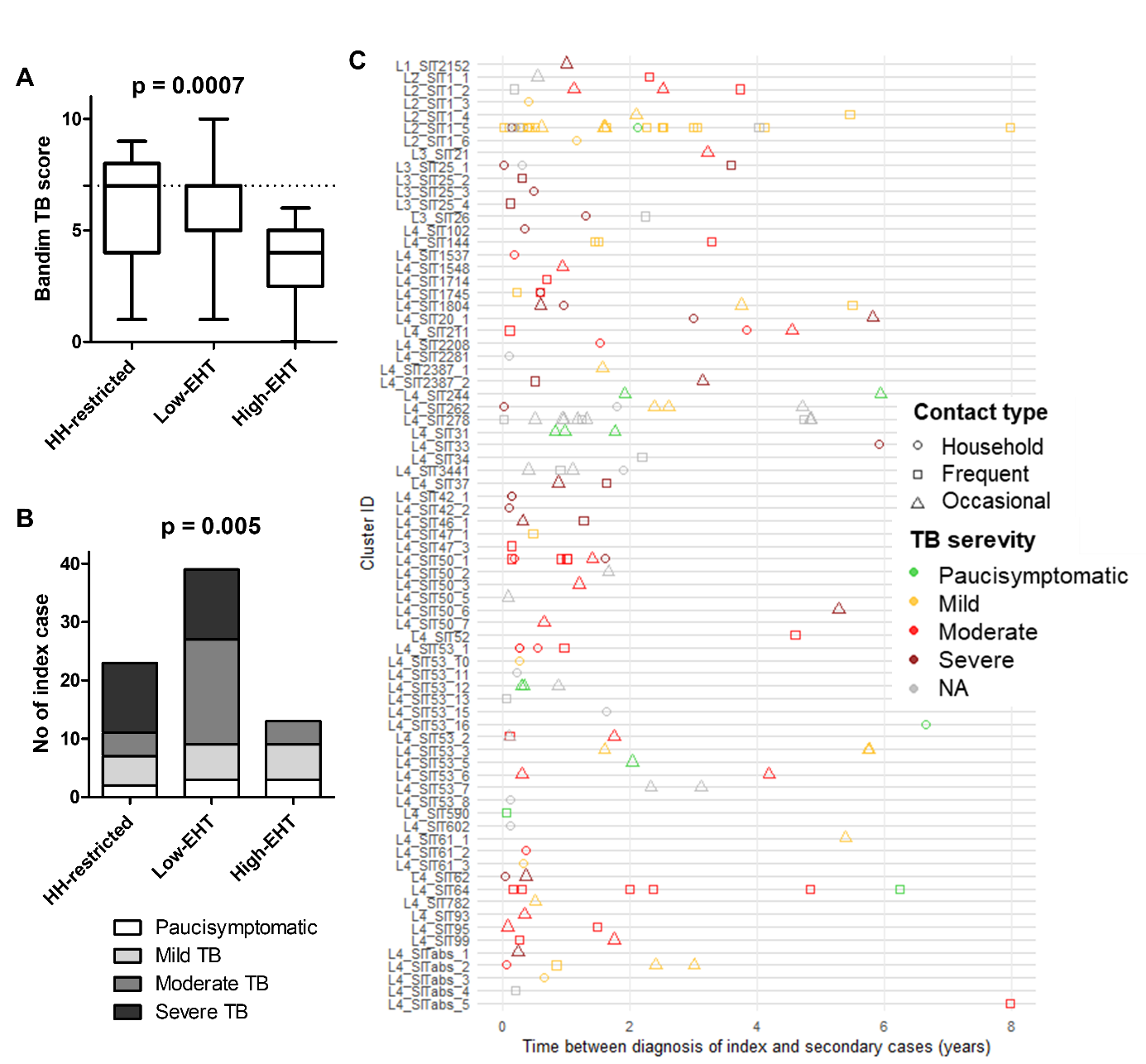

Figure S8: Trend in the Bandim TB score of index cases according to EHT level.

Index cases were stratified into three ordered categories according to their level of extra household transmission (EHT), by considering those transmitting TB: exclusively within household (HH-restricted), to at least one immunocompetent secondary case outside of household (Low-EHT) and to two or more immunocompetent secondary cases outside of household (High-EHT). (A) Trend in the median Bandim TB score of index cases according to EHT level, assessed using Jonckheere-Terpstra. (B) Trend in Bandim TB score of index cases, stratified as paucisymptomatic (≤2, white), mild TB (3-4, light grey), moderate TB (5-6, dark grey), severe TB (≥7, black) according to EHT level, assessed using Cochran-Armitage. (C) Each row represents a cluster with cluster ID indicating Mtb lineage and the shared international type (SIT) of the cluster determined by spoligotyping. The x-axis indicates the time between diagnosis of index and secondary cases (years), each symbol representing a transmission event. Symbol forms indicate epidemiological link between index and secondary cases: circle household contact, square frequent contact, triangle occasional contact. Symbol colours indicate the Bandim TB score of index case stratified as follow: paucisymptomatic (green), mild TB (yellow), moderate TB (red), severe TB (dark red) or not available data (NA, grey).

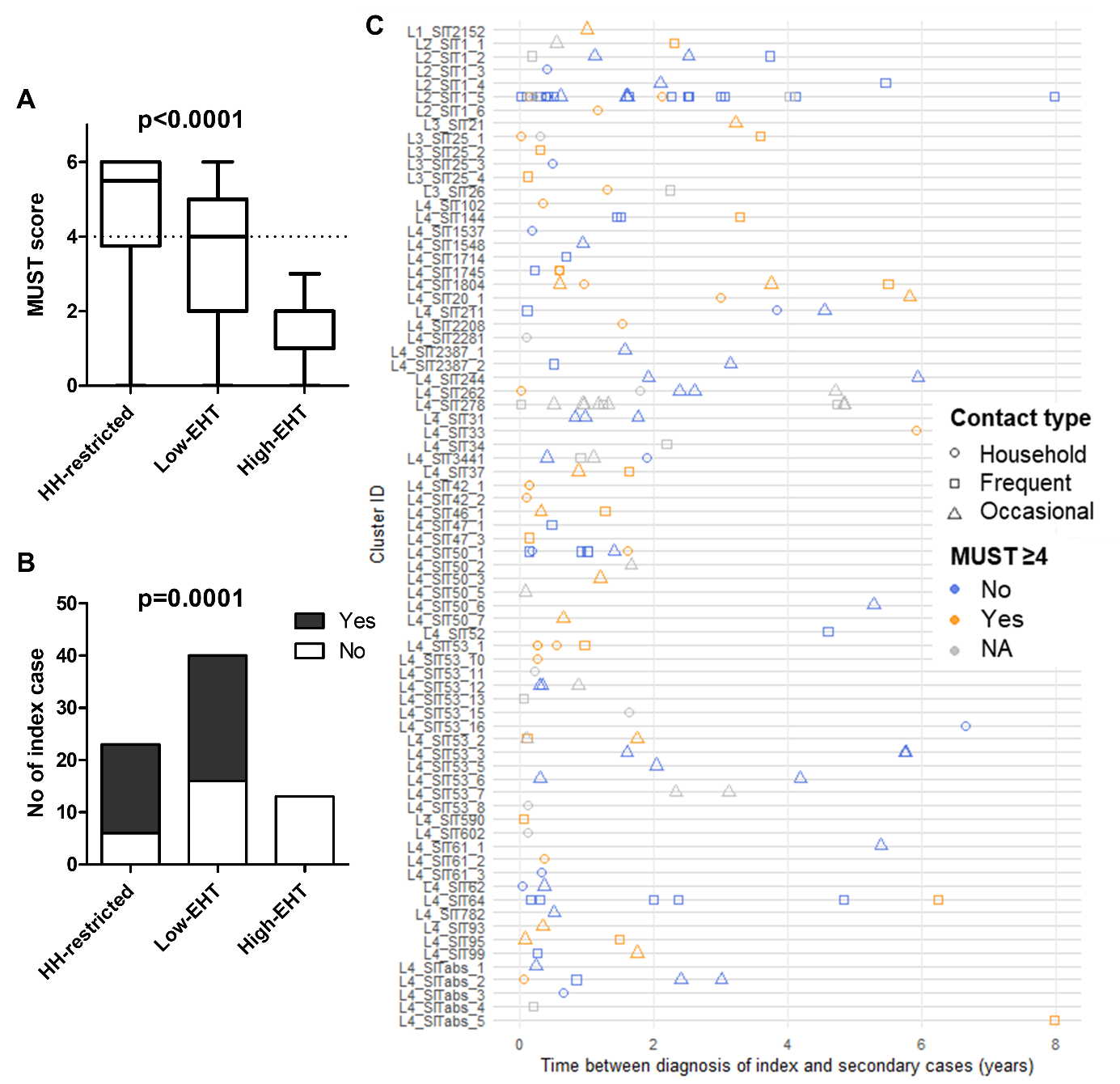

Figure S9: Trend in the nutritional status of index cases assessed by MUST according to EHT level.

Index cases were stratified into three ordered categories according to their level of extra household transmission (EHT), by considering those transmitting TB: exclusively within household (HH-restricted), to at least one immunocompetent secondary case outside of household (Low-EHT) and to two or more immunocompetent secondary cases outside of household (High-EHT). Nutritional status of index cases was assessed using the Malnutrition Universal Screening Tool (MUST). (A) Trend in the median MUST score in index cases according to EHT level, assessed using Jonckheere-Terpstra test. (B) Trend in the nutritional status of index cases, stratified by MUST score ≥4 (Yes: black; No; White) according to EHT level, assessed using Cochran-Armitage test. (C) Each row represents a cluster with cluster ID indicating Mtb lineage and the shared international type (SIT) of the cluster determined by spoligotyping. The x-axis indicates the time between diagnosis of index and secondary cases (years), each symbol representing a transmission event. Symbol forms indicate epidemiological link between index and secondary cases: circle household contact, square frequent contact, triangle occasional contact. Symbol colours indicate the nutritional status of the index case: MUST <4 (blue), MUST ≥4 (orange), or not available data (NA, grey).

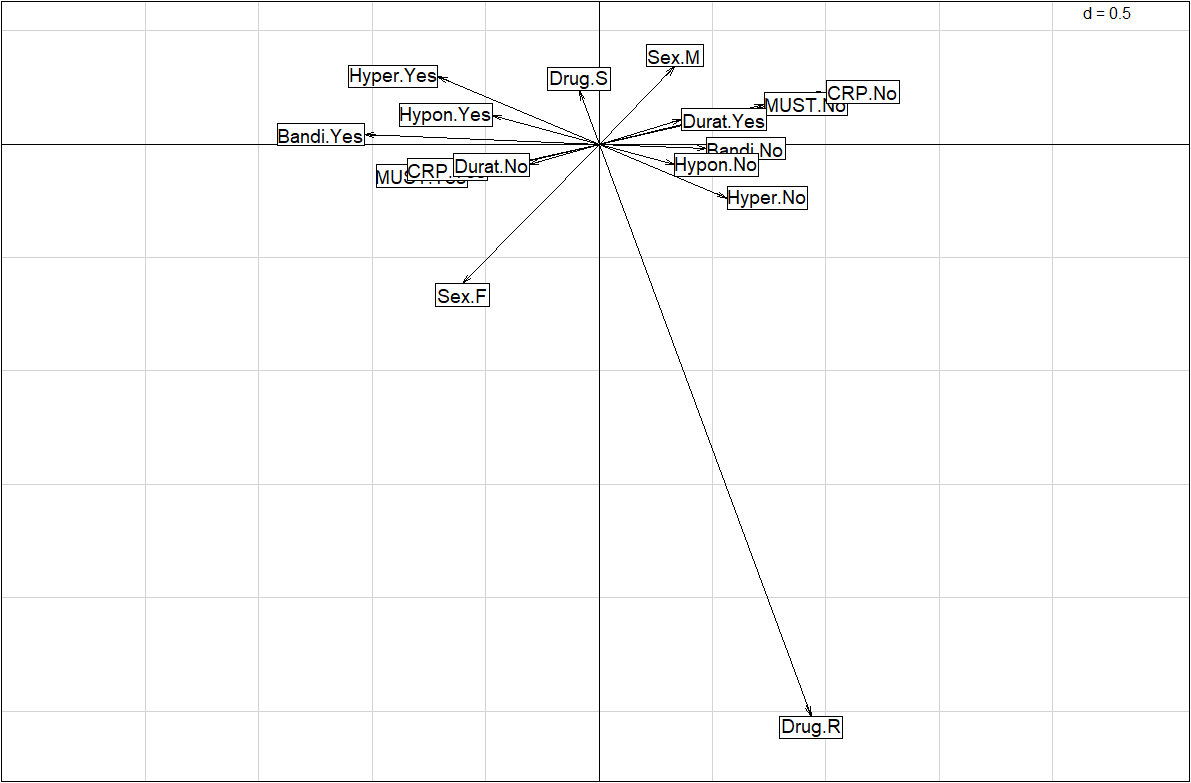

#### Figure S10: Variable contributions to EHT level identified by factor analysis of mixed data (FAMD).

Variables included in the analysis correspond to those with p<0.20 in univariable ordinal logistic regression, after exclusion of redundant or mathematically related variables. Observations with missing data for any included variable were excluded. Two-dimensional representation of variables projected onto the first and third principal axes of the FAMD, explaining 29.5% and 11.9% of total variance, respectively. Vectors indicate the direction and magnitude of the contribution of each variable to the principal axes shown in figure 4. Bandi: Bandim TB score ≥7, Yes or No; CRP: CRP ≥50 mg/mL, Yes or No; Drug: Mtb drug resistance, S (susceptible) or R (resistant to at least one first-line drug); Durat: Duration of symptoms before diagnosis ≥3 months, Yes or No; Hyper: Hyperneutrophilia, Yes or No; Hypon: Hyponatremia, Yes or No; MUST: MUST score ≥4, Yes or No; Sex: M (Male) or F (Female).
